# Rare-variant risk scores complement common-variant polygenic scores for disease risk prediction and stratification

**DOI:** 10.64898/2026.06.21.26356150

**Authors:** Menghan Qin, Bangsheng Wu, Liu Yang, Wei Cheng, Jianfeng Feng, Jintai Yu, Tian Ge, Weikang Gong

## Abstract

Polygenic risk scores (PRSs), which aggregate genetic effects across the genome, are typically constructed from common variants and therefore do not capture a substantial component of rare genetic variation. Using whole-genome sequencing data from the UK Biobank, we develop and benchmark rare-variant PRSs (rvPRSs) across 31 complex traits and 464 disease endpoints. Although rvPRSs provide only modest average improvements in population-level prediction beyond common-variant PRSs (cvPRSs), selected phenotypes show substantial discrimination driven by large-effect genes and rare-variant association signals not tagged by common-variant GWASs. At the individual level, rvPRSs identify largely nonoverlapping sets of individuals with extreme phenotypes or elevated disease risk compared with cvPRSs. These individuals are enriched for protein-truncating, damaging missense, or regulatory variants in biologically relevant genes, including those involved in lipid metabolism, liver function, cancer susceptibility, and cardiomyopathy. Survival analyses further show that rvPRSs stratify incident disease risk beyond cvPRSs over 15 years of follow-up. Together, these findings demonstrate that rvPRSs complement cvPRSs by enhancing tail-risk stratification and improving the biological interpretability of high-risk individuals.

## Introduction

Human complex traits and diseases are influenced by the cumulative effects of genetic variants spanning the full allele frequency spectrum. Polygenic risk scores (PRSs), which aggregate genetic effects across the genome, have shown promise for predicting disease risk, enabling early detection, facilitating patient stratification, and informing preventive strategies^1–5^. However, most existing PRSs are constructed from common genetic variants that are directly measured or tagged by genotyping arrays^3,6–9^, and therefore capture only a subset of the heritable genetic variation underlying complex traits and diseases.

The increasing availability of large-scale whole-exome and whole-genome sequencing (WGS) data has revealed a vast landscape of rare genetic variation^10,11^, which is hypothesized to contribute to the missing heritability^12–15^. Compared with common variants, disease-associated rare variants often have larger effect sizes and are enriched in functionally important genomic regions^16–19^, making them attractive targets for both risk prediction and therapeutic discovery. However, leveraging rare variants for polygenic prediction presents substantial methodological challenges. Their low frequencies and high dimensionality limit the effectiveness of established PRS construction methods developed for common variants, and best practices for aggregating rare variants across diverse functional annotations remain unclear. Although theoretical and empirical studies suggest that rare variants may explain a relatively modest proportion of phenotypic variance at the population level^12,20,21^, emerging evidence indicates that rare-variant PRSs (rvPRSs) may be particularly informative for identifying individuals at the extremes of the phenotypic distribution^22–24^, where clinical utility may be greatest. These considerations highlight the need for a systematic, large-scale evaluation of rvPRS construction strategies and predictive performance, including metrics that extend beyond conventional population-level measures to capture clinical relevance at the individual level.

In this study, building on recent advances in functional annotation of both coding and noncoding regions, including the STAAR framework^17,18,25^, we comprehensively evaluate linear and nonlinear approaches for constructing rvPRSs and assess their predictive performance across 31 complex traits and 464 disease endpoints using WGS data from the UK Biobank. We evaluate performance at both the population level and, critically, the individual level. Leveraging model-based attribution methods to pinpoint the genes and annotations contributing most strongly to prediction, we show that rvPRSs implicate large-effect genes and rare-variant association signals not captured by common-variant genome-wide association studies (GWASs). We further demonstrate that rvPRSs identify largely – and often completely – distinct sets of individuals with extreme phenotypes or elevated disease risk compared with common-variant PRSs (cvPRSs). These individuals are frequently carriers of high-impact rare coding variants or variants in regulatory elements, providing insight into underlying biological mechanisms. Finally, through survival analyses, we show that rvPRSs provide additional stratification of disease trajectories beyond cvPRSs over 15 years of follow-up.

Together, our results demonstrate that rvPRSs complement cvPRSs by capturing genetic signals from both coding and regulatory variation, enhancing the identification of high-risk individuals, and providing new insights into the genetic architecture of complex traits and diseases with important implications for clinical translation^4^.

## Results

### Overview of rvPRS construction

Large-scale whole-exome and whole-genome sequencing studies have shown that rare-variant associations are often enriched within specific functional annotations and allele frequency ranges. Using the STAARpipeline^17,18,25^, variants in protein-coding regions were annotated into four categories: (i) predicted loss-of-function (pLoF) variants, including stop-gain, stop-loss, frameshift, and splice-site variants; (ii) protein-truncation variants (PTVs); (iii) missense variants, further classified as disruptive or tolerated based on MetaSVM^26^ predictions; and (iv) synonymous variants. Using these annotations, variants were grouped into seven coding masks based on individual annotations and their combinations (e.g., pLoF, PTVs, missense, pLoF + disruptive missense; Methods). For noncoding regions, variants were organized into eight regulatory masks, including promoters and enhancers overlapping CAGE or DHS sites, untranslated regions (UTRs), upstream and downstream regions, and noncoding RNA (ncRNA) genes (Methods).

For each gene and minor allele frequency (MAF) threshold (MAF ≤1×10^−5^, ≤0.001, or ≤0.01), variants within each coding or noncoding mask were aggregated into burden scores, yielding 15 features per gene across 18,445 genes. These gene-level features were then used to construct coding and noncoding rvPRSs using both linear and nonlinear approaches. Specifically, we evaluated (i) linear or logistic regression; (ii) regularized generalized linear models (GLMNET)^27^; (iii) multi-layer perceptrons (MLPs); (iv) light gradient boosting machines (LightGBM)^28,29^; and (v) RVTrans, a Transformer-based model^30,31^ tailored for rvPRS construction (Methods).

We randomly partitioned 337,988 unrelated UK Biobank participants of European ancestry into training (80%), validation (10%), and testing (10%) sets. Models were trained in the training set, with hyperparameters and the optimal MAF threshold selected in the validation set. Final performance of coding and noncoding rvPRSs was evaluated in the independent testing set. An overview of the study design is summarized in Figure 1.

**Figure 1:**
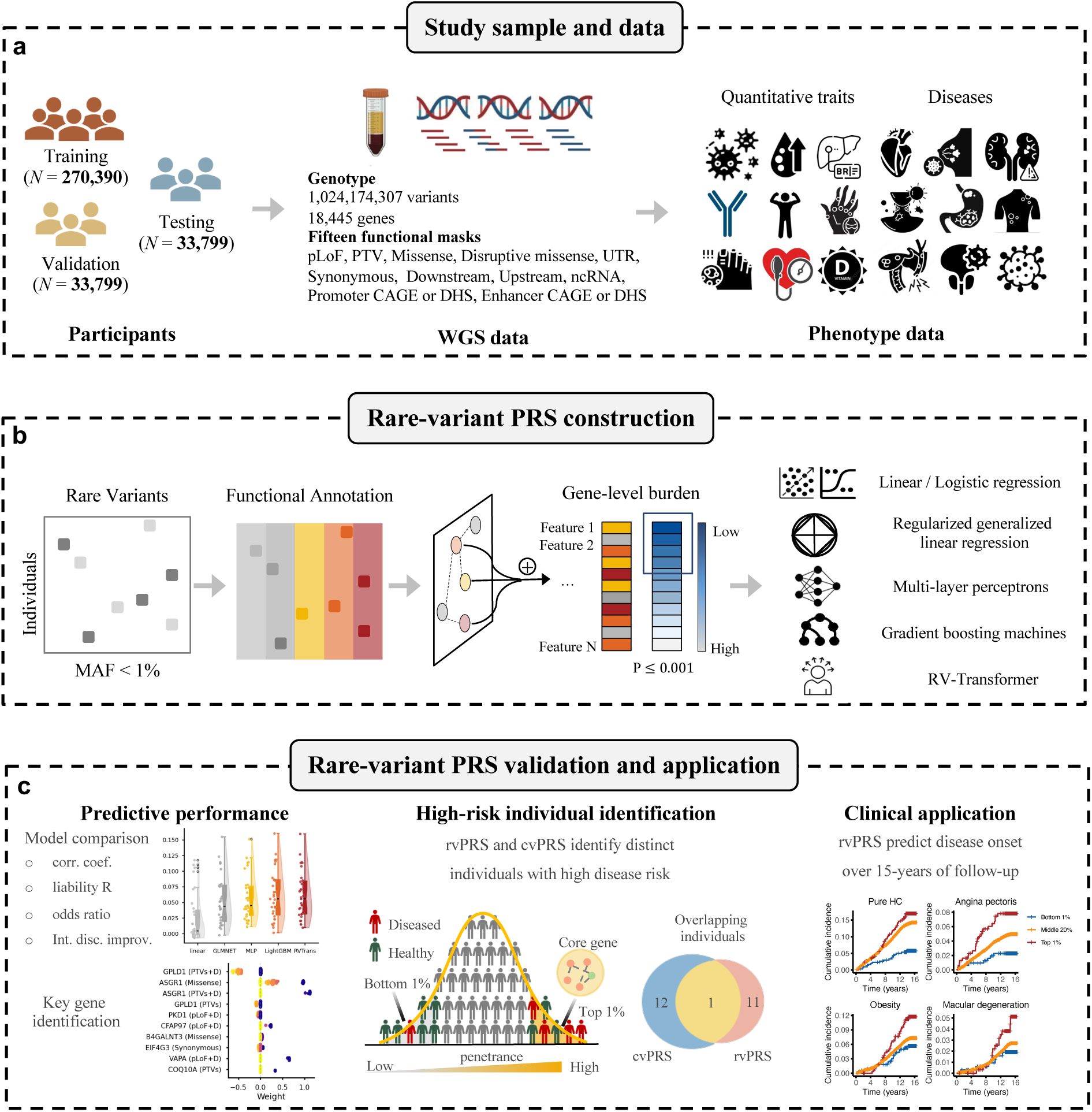
Overview of the study design. **a,** Study sample and data. Unrelated UK Biobank participants of European ancestry were partitioned into training (80%), validation (10%), and testing (10%) sets. Whole-genome sequencing variants were annotated into functional categories across coding and noncoding regions. Predictive performance of common- and rare-variant polygenic risk scores (cvPRSs and rvPRSs) was evaluated across 31 complex traits and 464 disease endpoints. **b,** Construction of rvPRSs. For each gene and minor allele frequency (MAF) threshold, variants within each coding or noncoding mask were aggregated into burden scores, which served as input features for five linear and nonlinear prediction models. **c,** Evaluation of rvPRSs. Predictive performance was assessed at both the population level, using multiple metrics, and the individual level, including identification of high-risk individuals and stratification of disease trajectories over 15 years of follow-up.

### Population-level predictive performance of rvPRSs across complex traits and diseases

We first evaluated the population-level predictive performance of rvPRSs generated by different methods, benchmarking them against cvPRSs constructed using PRS-CS^3,6^ and trained in the same dataset. Analyses were conducted for 31 quantitative traits (Supplementary Table 1) and 119 diseases with prevalence >5% (Supplementary Table 2). Predictive performance was assessed using three complementary metrics: (i) the correlation *R* between predicted and observed phenotypes, with disease endpoints analyzed on the liability scale; (ii) the odds ratio (OR) comparing individuals in the top 1% of the rvPRS distribution with the remainder of the sample, with individuals in the top 1% of the observed phenotype distribution treated as cases for quantitative traits; and (iii) the integrated discrimination improvement (IDI) obtained by adding rvPRSs to a model already including cvPRSs, using the top 1% of the rvPRS distribution as the risk threshold. All evaluation models adjusted for age, sex, and top 10 genetic principal components (PCs). To isolate the incremental contribution of rvPRSs, we additionally included cvPRSs as covariates when computing *R* and OR for rvPRSs.

For quantitative traits, an MAF cutoff of 0.01 produced the most predictive coding rvPRSs in the majority of cases (96.8%) compared with more stringent thresholds (Figure 2a; Supplementary Table 3). After selecting the optimal MAF threshold, nonlinear machine learning models generally outperformed linear approaches, with LightGBM and RVTrans often achieving the highest accuracy across metrics (Figure 2c; Supplementary Table 3). Nevertheless, the incremental contribution of rvPRSs to population-level prediction was modest relative to cvPRSs, which showed stronger performance relative to a covariate-only model (median *R =* 0.276, median OR = 4.63, median IDI = 6.7×10^−3^; Supplementary Table 3), consistent with the low allele frequencies and limited population-level variance explained by rare variants.

**Figure 2:**
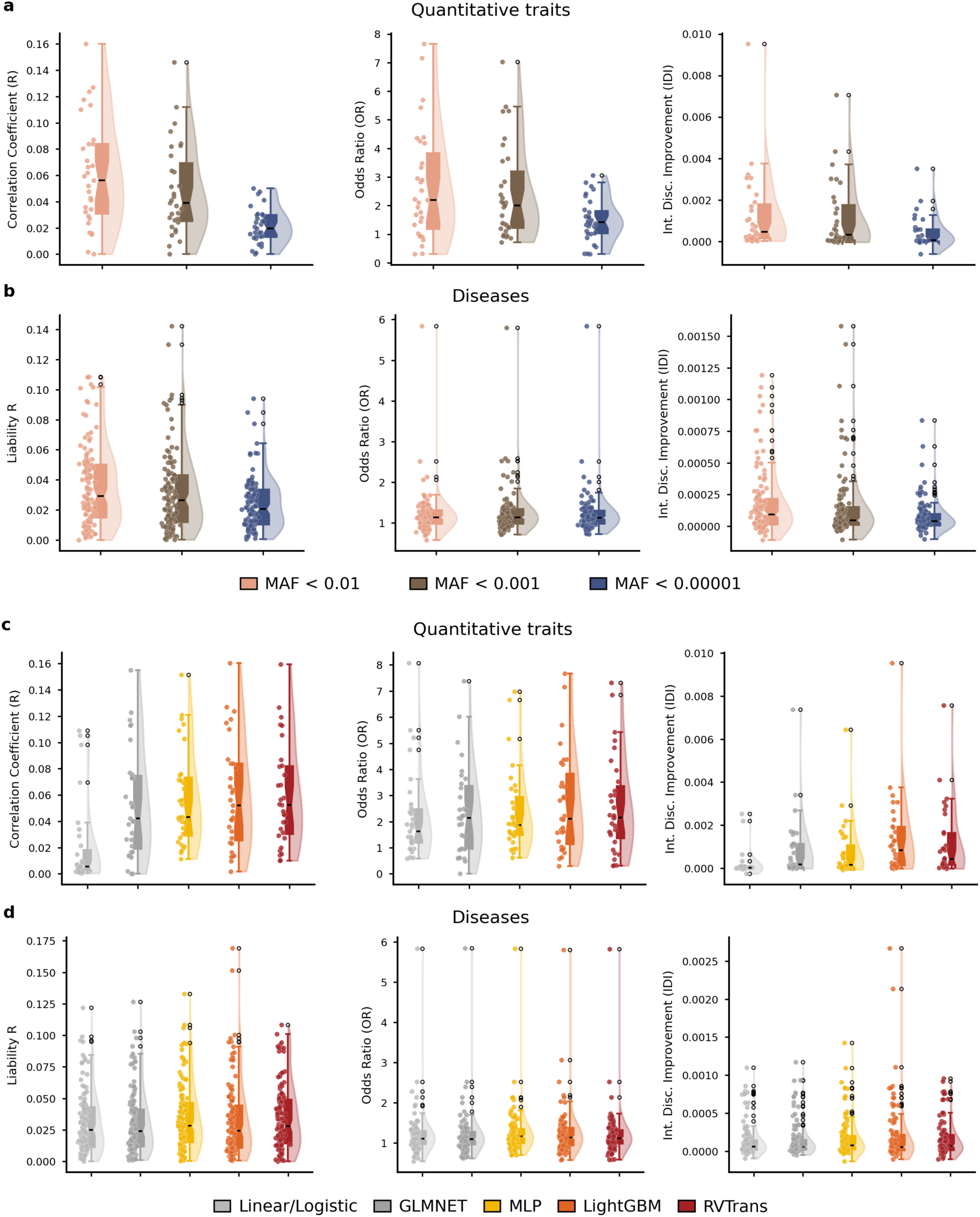
Population-level predictive performance of coding rare-variant polygenic risk scores across complex traits and diseases. **a,** Incremental predictive performance of rvPRSs beyond cvPRSs for 31 quantitative traits, evaluated using three metrics: (i) the correlation *R* between predicted and observed phenotypes, (ii) the odds ratio (OR) comparing individuals in the top 1% of the rvPRS distribution with the remainder of the sample, and (iii) the integrated discrimination improvement (IDI), across three different minor allele frequency (MAF) cutoffs. **b,** Incremental predictive performance of rvPRSs beyond cvPRSs for 119 diseases with prevalence >5%, evaluated across three MAF cutoffs. **c,** Incremental predictive performance of rvPRSs beyond cvPRSs for 31 quantitative traits across five prediction models. **d,** Incremental predictive performance of rvPRSs beyond cvPRSs for 119 diseases with prevalence >5% across five prediction models. Across all panels, each data point represents the optimal result for a given trait or disease, selected across prediction models (for panels **a**-**b**) or across MAF cutoffs (for panels **c**-**d**). The center line of each box plot denotes the median, and box limits indicate the interquartile range.

For binary disease endpoints, the optimal MAF cutoff for coding rvPRSs varied across phenotypes, likely reflecting differences in genetic architecture and statistical power (Figure 2b; Supplementary Table 4). Predictive performance was broadly similar across machine learning models, with LightGBM and RVTrans showing modest improvements over alternative approaches (Figure 2d; Supplementary Table 4). As with quantitative traits, rvPRSs contributed only a small fraction of predictive power relative to cvPRSs, which showed stronger performance relative to a covariate-only model (median *R =* 0.248, median OR = 1.73, median IDI = 4.6×10^−3^; Supplementary Table 4).

Noncoding rvPRSs exhibited patterns similar to coding rvPRSs but had lower predictive accuracy overall (Supplementary Figure 1; Supplementary Tables 5-6), consistent with the more indirect functional impact of most noncoding variants. Sensitivity analyses using alternative risk thresholds, defined as the top 2% or 5% of the rvPRS distribution, yielded similar conclusions (Supplementary Figure 2; Supplementary Tables 3-6).

Overall, although integrating gene- and annotation-level burden scores enabled statistically significant rvPRS predictions for many traits and diseases, their contribution to population-level prediction remained modest relative to cvPRSs. Given the stronger performance of LightGBM and RVTrans compared with alternative methods, together with the computational efficiency of LightGBM, we extended LightGBM-based rvPRS analyses to an additional 345 disease endpoints with lower prevalence (Supplementary Tables 7-8) and focus on these results in subsequent sections.

### Large-effect genes drive strong rvPRS performance for selected traits and diseases

Although the overall contribution of rvPRSs to population-level prediction was modest, several traits and diseases showed substantial rare-variant contributions, with rvPRS performance comparable to, and in some cases exceeding, that of cvPRSs. We highlight representative examples using LightGBM.

Among quantitative traits, alkaline phosphatase (ALP) showed the strongest rvPRS predictive performance in both coding (Figure 3a; *R* = 0.160; OR = 4.39, 95% CI 2.52-7.64; IDI = 3.1×10^−3^) and noncoding regions (Figure 3c; *R* = 0.115; OR = 7.65, 95% CI 4.93-11.86; IDI = 7.7×10^−3^), indicating a substantial contribution of rare variation to circulating ALP levels. SHapley Additive exPlanations (SHAP) based feature importance analyses^32,33^ identified PTV and disruptive missense masks in *GPLD1* and *ASGR1* as major contributors to the coding rvPRS (Figure 3a; Supplementary Table 9). Both genes are known to influence circulating ALP through liver-specific pathways that regulate the release and clearance of glycoproteins^34^. Independent external WGS association analyses in the *All of Us* (AoU) Research Program^35,36^ confirmed significant rare-variant burden associations with consistent directions of effect (*GPLD1*: *p* = 7.95×10^-21^; *ASGR1*: *p* = 3.81×10^-14^). In noncoding regions, *ASGR2*^37^ and *EIF5A* emerged as top contributors (Figure 3c; Supplementary Table 9), with the latter potentially acting through regulatory effects on liver-expressed genes or protein synthesis pathways affecting hepatocyte function^38,39^.

**Figure 3:**
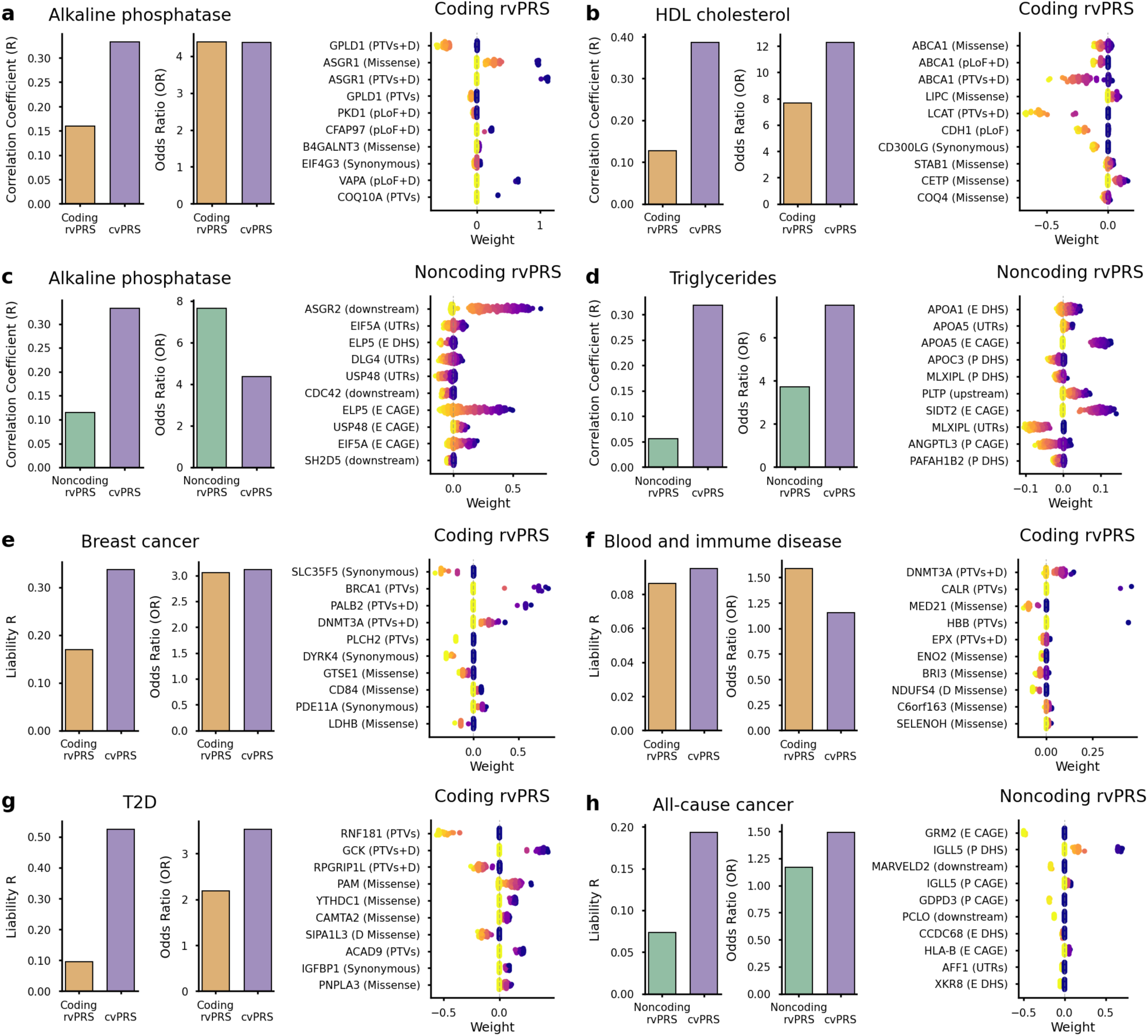
Large-effect genes drive predictive performance of rare-variant polygenic risk scores for selected traits and diseases. For each trait or disease, the left panel shows prediction accuracy and risk discrimination for rvPRSs and cvPRSs, with discrimination quantified by the odds ratio comparing individuals in the top 1% of the corresponding PRS distribution with the reminder of the sample. The right panel shows the genes and functional annotations with the strongest contributions to rvPRS prediction, based on SHapley Additive exPlanations (SHAP) feature importance analysis implemented in the LightGBM models.

For high-density lipoprotein (HDL) cholesterol, the coding rvPRS also showed strong predictive performance (Figure 3b; *R* = 0.127; OR = 7.66, 95% CI 4.71-12.46; IDI = 9.5×10^−3^), driven by established lipid genes including *ABCA1*^40^ (Supplementary Table 9; AoU *p* = 5.38×10^-21^) and *LIPC*^41^ (Supplementary Table 9; AoU *p* = 4.60×10^-3^). In parallel, the noncoding rvPRS contributed substantially to triglyceride levels (Figure 3d; *R* = 0.056; OR = 3.70, 95% CI 2.06-6.64; IDI = 2.8×10^−3^), with major contributions from apolipoprotein (*APO*) genes^42^, including *APOA1*^43^, *APOC3*^44^, and *APOA5*^45^, which play central roles in lipoprotein metabolism.

Similar large-effect gene contributions were observed for disease endpoints. For breast cancer, the coding rvPRS achieved strong risk discrimination (Figure 3e; liability *R* = 0.169; OR = 3.06, 95% CI 2.03-4.61; IDI = 2.7×10^−3^). Feature importance analyses highlighted established susceptibility genes, including *BRCA1* (Supplementary Table 10; AoU *p* = 9.47×10^-41^), for which pathogenic variants confer a lifetime breast cancer risk of more than 50%^46,47^; *PALB2* (Supplementary Table 10; AoU *p* = 6.48×10^-9^), a tumor suppressor gene that drastically increases breast cancer risk^48^; and *DNMT3A*, which modulates DNA methylation programs in breast cancer cells^49^.

For blood and immune disease, the predictive performance of the coding rvPRS (Figure 3f; liability *R* = 0.086; OR = 1.59, 95% CI 1.21-2.09 IDI = 8.3×10^−4^) was largely driven by PTVs and disruptive missense variants in *DNMT3A*, one of the most frequently mutated genes in age-related clonal hematopoiesis, with alterations implicated in hematological malignancies and immune dysregulation^50^. Additional contributions came from PTVs in *CALR*, a major driver gene in chronic myeloid neoplasms, including essential thrombocythemia and myelofibrosis^51^. The coding rvPRS also showed strong predictive performance for type 2 diabetes (Figure 3g; liability *R* = 0.095; OR = 2.19, 95% CI 1.60-2.99; IDI = 8.6×10^−4^), implicating the established diabetes gene *GCK*^52^ (Supplementary Table 10; AoU *p* = 7.63×10^-5^), along with *RNF181*, which has been previously linked to core metabolic regulatory pathways^53^.

In noncoding regions, the rvPRS for all-cause cancer (Figure 3h; liability *R* = 0.074; OR = 1.17, 95% CI 0.91-1.51; IDI = 5.3×10^−4^) was driven by genes with broad oncogenic relevance, including *GRM2*^54^, associated with DNA methylation and glutamate receptor regulation; *IGLL5*^55^, involved in tumor immune infiltration; *MARVELD2*^56^, linked to poor prognosis in breast cancer; and *GDPD3*^57^, which regulates lysophosphatidic acid-mediated tumor progression. These findings suggest that rare regulatory variation may contribute to cancer susceptibility through diverse molecular pathways.

Additional examples for both quantitative traits and disease endpoints are shown in Supplementary Figures 3-4. In summary, these results demonstrate that, for selected complex traits and diseases, large-effect genes not captured by cvPRSs can substantially contribute to rvPRS performance, enabling rvPRSs to achieve comparable – and in some cases superior – prediction accuracy and risk discrimination relative to cvPRSs.

### rvPRSs and cvPRSs identify distinct individuals with extreme phenotypes

Consistent with the observation that population-level rvPRS performance is largely driven by rare-variant association signals not captured by common-variant GWASs, we hypothesized that rvPRSs would identify distinct sets of individuals with extreme phenotypes compared with those identified by cvPRSs.

Using alkaline phosphatase (ALP) as an example, among individuals ranked in the top 1% by the coding rvPRS or cvPRS, 14 from each group had ALP levels in the top 1% of the testing sample, with only one individual shared between the two PRSs (Figure 4a; Supplementary Table 11). As expected, individuals identified by the coding rvPRS were strongly enriched for carriers of PTVs and missense variants in genes driving rvPRS performance (Figure 3a). Among individuals in the top 1% of the coding rvPRS distribution, more than half carried missense variants in *ASGR1* (AoU *p* = 3.81×10^−14^), which has been associated with an approximately 50% increase in plasma ALP levels^34,58^. In contrast, few individuals identified by the cvPRS carried missense variants in this gene (Figure 4a; Supplementary Table 11). The noncoding rvPRS for ALP identified substantially more individuals with ALP levels in the top 1% than the cvPRS (Figure 4a; Supplementary Table 11), consistent with its strong discrimination (Figure 3c). Individuals identified by the noncoding rvPRS were strongly enriched for variants near genes driving the rvPRS signal, including regulatory regions linked to *ELP5* and *ASGR2*^37^, which were not captured by the cvPRS (Figures 3c & 4a; Supplementary Table 11).

**Figure 4:**
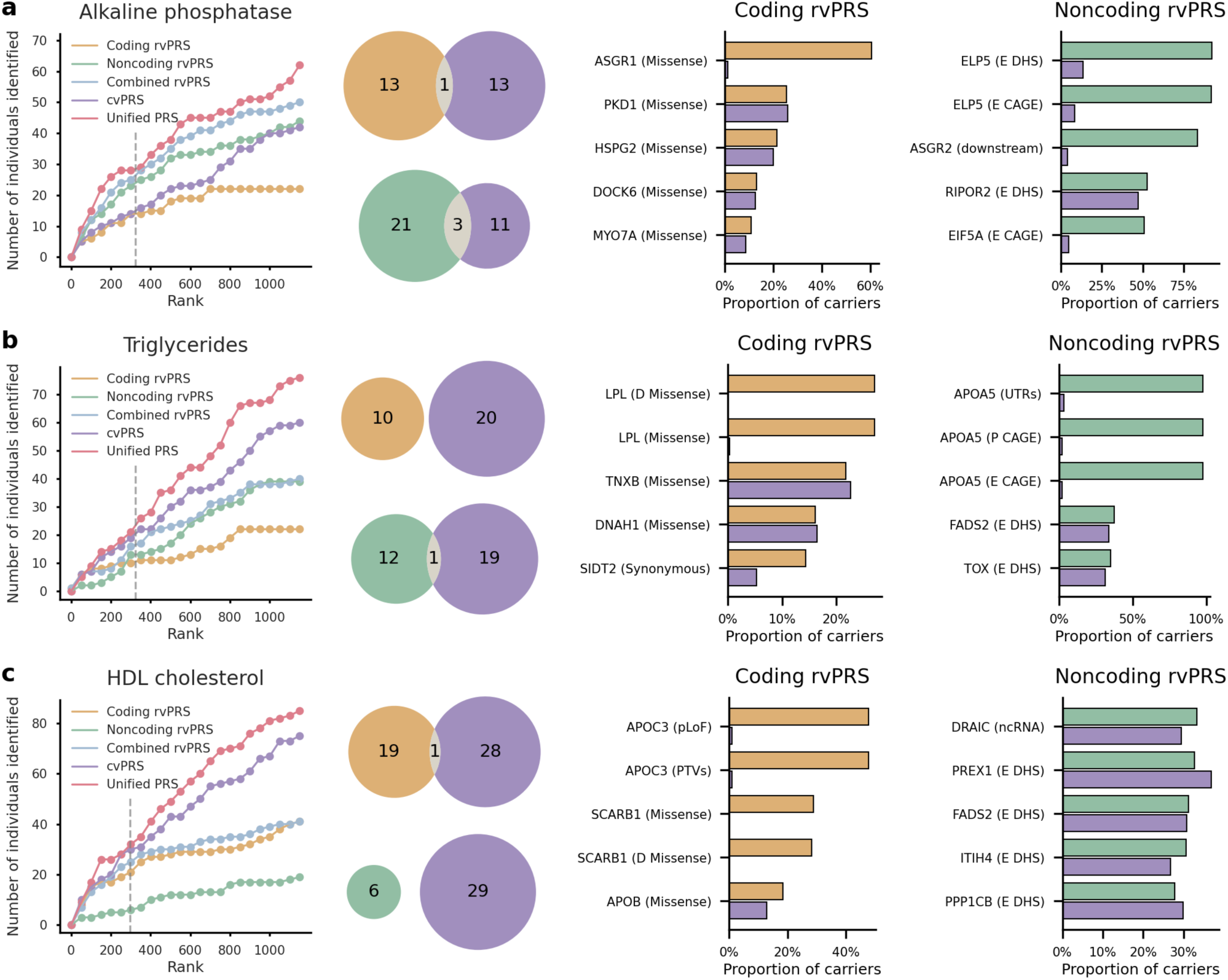
Identification of individuals with extreme phenotypes using rare-variant, common-variant, and integrated polygenic risk scores for selected complex traits. For each trait, the left panel shows the number of individuals whose phenotype falls within the top 1% of the testing sample as a function of PRS rank for the coding rvPRS, noncoding rvPRS, combined rvPRS, cvPRS, and unified PRS integrating rvPRSs and cvPRSs. The vertical dashed gray line indicates the top 1% threshold of the PRS distribution. The middle panel presents Venn diagrams showing the number of individuals with phenotypes in the top 1% of the testing sample who were identified by the coding rvPRS, noncoding rvPRS, and cvPRS, along with their overlaps. The right panel shows genes and functional annotations enriched among individuals identified by the coding rvPRS, noncoding rvPRS, and cvPRS.

We next combined coding and noncoding rvPRSs into a single rvPRS using a linear model and further integrated this combined rvPRS with the cvPRS into a unified PRS. This integrated model leveraged complementary signals across scores and identified more individuals with extreme phenotypes than any single PRS alone (Figure 4a; Supplementary Table 12).

As a second example, rvPRSs and cvPRSs identified highly distinct sets of individuals with extreme triglyceride levels (Figure 4b; Supplementary Table 11). The coding rvPRS showed unique enrichment for missense variants in *LPL* (AoU *p* = 1.18×10^−5^), which encodes a key enzyme responsible for triglyceride hydrolysis in circulation^59^. Individuals identified by the noncoding rvPRS were almost exclusively carriers of variants in regulatory regions near *APOA5*, consistent with its central role in triglyceride metabolism and lipoprotein processing^60^; these signals were largely absent from the cvPRS (Figure 4b; Supplementary Table 11). The unified PRS integrating the rvPRS and cvPRS outperformed each individual score (Figure 4b; Supplementary Table 12).

A similar pattern was observed for HDL cholesterol (Figure 4c; Supplementary Tables 11-12). Individuals identified by the coding rvPRS were uniquely enriched for PTVs and pLoF variants in *APOC3* (AoU *p* = 1.71×10^−11^), a key regulator of lipoprotein metabolism^61^, as well as missense variants in *SCARB1* (AoU *p* = 1.79×10^−13^), which encodes the primary hepatic receptor for HDL. Individuals identified by the noncoding rvPRS showed enrichment for variants in enhancer regions overlapping DHS sites near *FADS2*, which encodes desaturase enzymes involved in polyunsaturated fatty acid metabolism^62^, and *ITIH4*, previously implicated in plasma cholesterol regulation^63^. Notably, although the noncoding rvPRS and cvPRS identified completely nonoverlapping sets of individuals with high HDL cholesterol, both sets were enriched for similar regulatory elements, suggesting convergence of rare- and common-variant signals on shared biological pathways (Figure 4c; Supplementary Table 11).

Additional examples are shown in Supplementary Figure 5. Together, these results indicate that rvPRSs can identify additional individuals with extreme phenotypes who are not captured by cvPRSs. These individuals are often enriched for rare coding and regulatory variants in large-effect genes, providing insight into underlying biological pathways and disease mechanisms.

### rvPRSs and cvPRSs identify distinct individuals with high disease risk

We extended these analyses to disease endpoints. Figure 5 presents illustrative examples in which rvPRSs, using the top 1% of the score distribution as the cutoff, often identified largely or completely distinct sets of cases, with case counts comparable to or greater than those identified by the top 1% of the cvPRS distribution.

**Figure 5:**
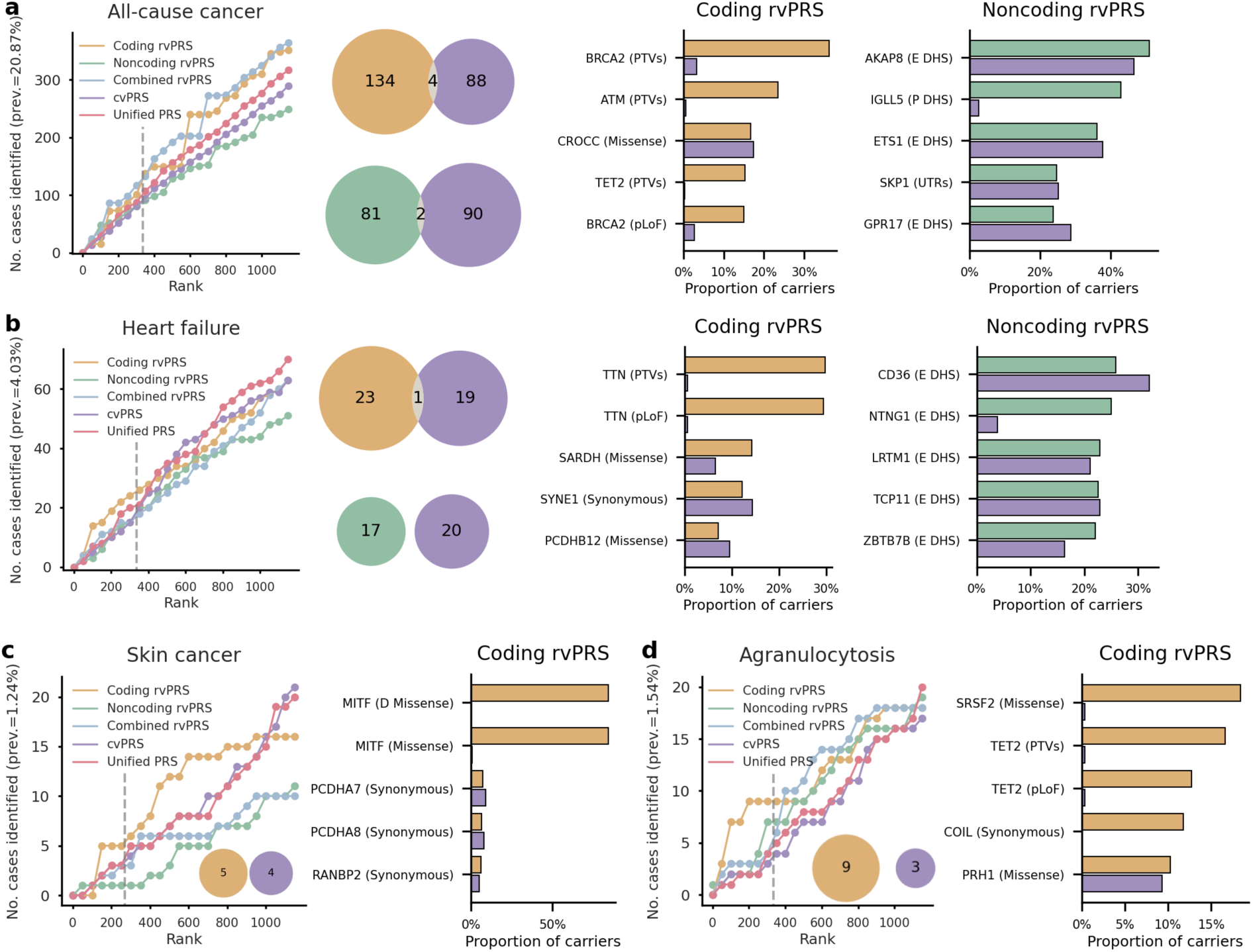
Identification of high-risk individuals for selected diseases using rare-variant, common-variant, and integrated polygenic risk scores. **a-b,** The left panel shows the number of cases as a function of PRS rank for the coding rvPRS, noncoding rvPRS, combined rvPRS, cvPRS, and unified PRS integrating rvPRSs and cvPRSs. The vertical dashed gray line indicates the top 1% threshold of the PRS distribution. The middle panel presents Venn diagrams showing the number of cases identified by the coding rvPRS, noncoding rvPRS, and cvPRS, along with their overlaps. The right panel shows genes and functional annotations enriched among individuals identified by the coding rvPRS, noncoding rvPRS, and cvPRS. **c-d,** The left panel shows the number of cases as a function of PRS rank for different PRSs. Embedded Venn diagrams show the number of cases identified by the coding rvPRS and cvPRS. The right panel shows genes and functional annotations enriched among individuals identified by the coding rvPRS and cvPRS.

For all-cause cancer, coding and noncoding rvPRSs captured pathogenic signals across diverse cancer types. The coding rvPRS uniquely identified individuals enriched for PTVs in *BRCA2*^64^ (AoU *p* = 1.84×10^−31^) and *ATM*^65^ (AoU *p* = 6.17×10^−7^), both of which are known to substantially increase lifetime risk of breast, pancreatic, prostate, and ovarian cancers, as well as *TET2* (AoU *p* = 2.56×10^−2^), whose deficiency has been linked to hematopoietic cancers and solid malignancies^66^. The noncoding rvPRS identified individuals enriched for variants in regulatory elements near *AKAP8*^67,68^, *IGLL5*^55^, and *ETS1*^69,70^, genes implicated across a broad spectrum of cancers, including breast, lung, renal, and cervical cancers (Figure 5a; Supplementary Table 13). A combined rvPRS integrating coding and noncoding rare-variant signals identified the largest number of cases, highlighting the complementary effects of coding and regulatory variation on cancer susceptibility (Figure 5a; Supplementary Table 14).

A similar pattern was observed for heart failure (Figure 5b; Supplementary Tables 13-14). Individuals identified by the coding rvPRS were uniquely enriched for PTVs and pLoF variants in *TTN* (AoU *p* = 9.12×10^−11^), the most common genetic cause of familial dilated cardiomyopathy^71^, as well as missense variants in *SARDH*, a gene involved in mitochondrial metabolism^72^. Individuals identified by the noncoding rvPRS showed enrichment for variants in enhancer regions overlapping DHS sites near *CD36*, which encodes a fatty acid transporter that facilitates long-chain fatty acid uptake in cardiomyocytes^73^, and *LRTM1*, a regulator of intrinsic myocardial regeneration whose loss promotes cardiomyocyte proliferation^74^ (Figure 5b; Supplementary Table 13). Notably, individuals identified by the noncoding rvPRS showed unique enrichment of variants in enhancer regions near *NTNG1* compared with those identified by the cvPRS, while converging on other signals (Figure 5b; Supplementary Table 13). Although *NTNG1* has been associated with vascular health^75^ and diabetes^76^, direct evidence linking it to heart failure remains limited.

Finally, we examined two diseases with biologically informative signals but relatively lower prevalence. Coding rvPRSs for skin cancer and agranulocytosis identified distinct – and often more – cases than cvPRSs (Figure 5c-d; Supplementary Table 13). For skin cancer, the predictive signal of the coding rvPRS was primarily driven by missense mutations in *MITF*, a master regulator of melanocyte development and a major risk factor for melanoma^77^ (Figure 5c; Supplementary Table 13). For agranulocytosis, the coding rvPRS highlighted carriers of missense variants in *SRSF2*^78^, as well as PTVs and pLoF variants in *TET2*^79^ (Figure 5d; Supplementary Table 13). These genes implicate myeloid regulatory pathways and neutrophil differentiation, and the corresponding signals were largely absent among individuals identified by the cvPRS (Figure 5d; Supplementary Table 13).

Additional examples are shown in Supplementary Figures 6-7. Collectively, these findings indicate that rvPRSs can identify high-risk individuals not captured by cvPRSs through both distinct and partially overlapping genetic mechanisms, potentially informing disease subtypes as well as strategies for prevention and treatment.

### rvPRSs predict disease onset over 15 years of follow-up

Lastly, we performed survival analyses to evaluate associations between rvPRSs and the onset of 464 diseases over 15 years of follow-up (Supplementary Tables 15-16). Among individuals who were disease-free at baseline, LightGBM-derived coding rvPRSs were nominally significant predictors (*p* <0.05) of disease onset in the testing set for 76 diseases, based on Cox proportional hazards models adjusting for age, birth year, sex, top genetic PCs, and cvPRSs (Supplementary Table 15). Notable examples included all-cause cancer (hazard ratio [HR] = 1.08, 95% CI 1.06-1.11; *p* = 8.75×10^−13^), cardiomyopathy (HR = 1.31, 95% CI 1.16-1.47; *p* = 8.32×10^−6^), breast cancer (HR = 1.13, 95% CI 1.07-1.19; *p* = 1.32×10^−5^), and acute renal failure (HR = 1.10, 95% CI 1.05-1.15; *p* = 5.43×10^−5^) (Figure 6a). Noncoding rvPRSs also contributed to prediction of onset for 36 diseases (*p* <0.05; Supplementary Table 16).

**Figure 6:**
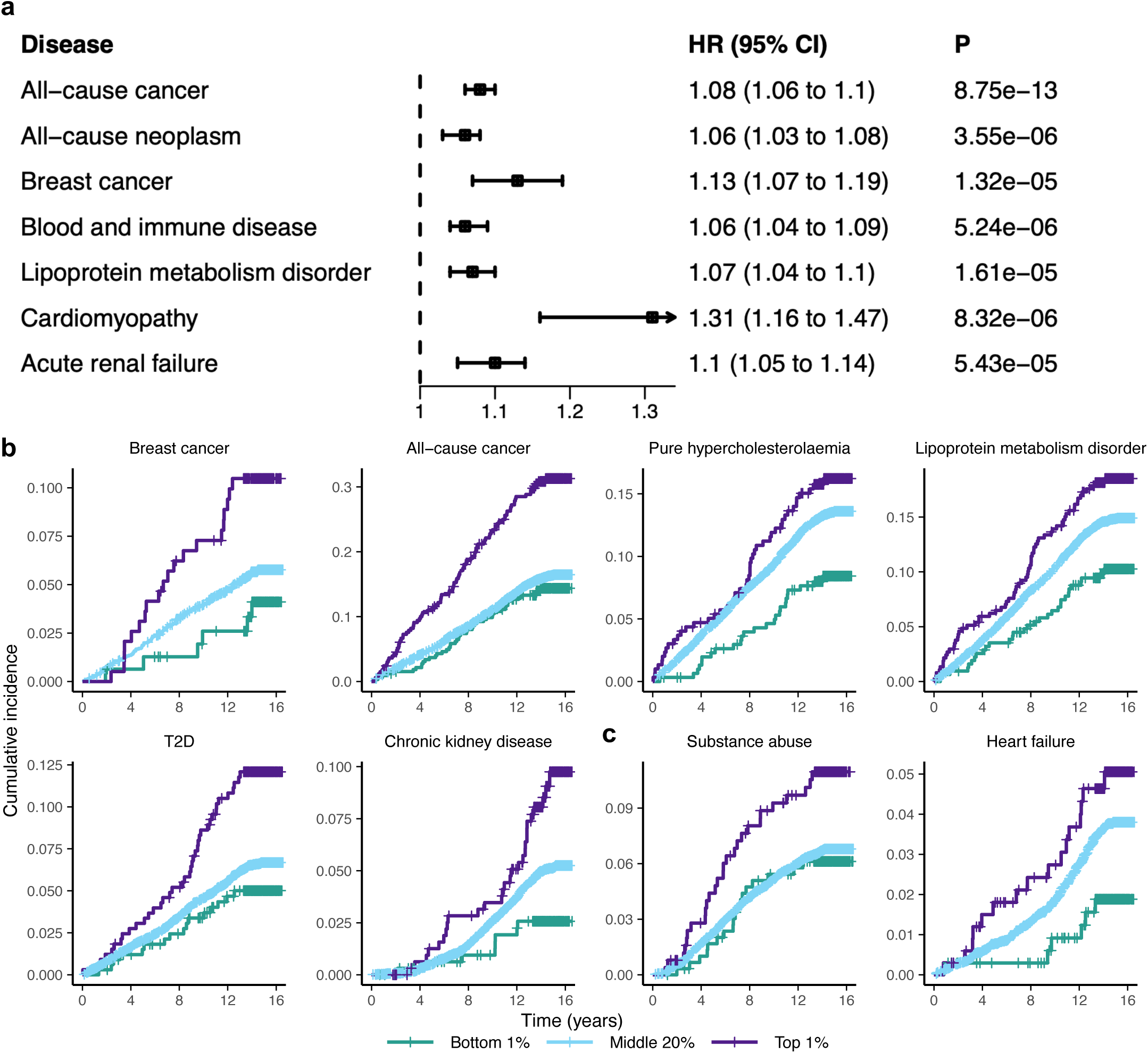
Rare-variant polygenic risk scores predict disease onset and stratify disease trajectories over 15 years of follow-up. **a,** Hazard ratios for LightGBM-derived coding rvPRSs for the onset of selected diseases, estimated using Cox proportional hazards models adjusting for age, birth year, sex, genetic PCs, and cvPRSs. **b,** Kaplan-Meier curves for the onset of representative diseases, stratified by the top 1%, middle 20%, and bottom 1% of the coding rvPRS distribution. **c,** Kaplan-Meier curves for the onset of representative diseases, stratified by the top 1%, middle 20%, and bottom 1% of the noncoding rvPRS distribution.

We further stratified participants into the top 1%, middle 20%, and bottom 1% of the rvPRS distributions. Kaplan-Meier curves for eight representative diseases (Figure 6b-c) showed that individuals in the top 1% of the rvPRS distribution had higher rates of incident disease than those at average or low rare-variant risk. For instance, the coding rvPRS for chronic kidney disease (top vs. bottom HR = 3.21, 95% CI = 1.41-7.29; *p* = 0.0054; Supplementary Table 17) and the noncoding rvPRS for heart failure (top vs. bottom HR = 3.03, 95% CI = 1.16-7.88; *p* = 0.023; Supplementary Table 17) showed approximately three-fold higher risk in the top 1% relative to the bottom 1% (Figure 6b-c). Additional examples are shown in Supplementary Figures 8-9. These results indicate that rvPRSs provide additional predictive information for disease onset beyond that captured by cvPRSs.

## Discussion

In this study, we developed and systematically benchmarked rvPRSs using whole-genome sequencing data from the UK Biobank across 31 complex traits and 464 disease endpoints. Although the average contribution of rvPRSs to population-level prediction was modest relative to cvPRSs, rvPRSs achieved comparable – or even higher – predictive performance for selected traits and diseases. These gains were primarily driven by association signals from large-effect genes not captured by common-variant GWASs. Importantly, at the individual level, rvPRSs identified largely nonoverlapping sets of individuals with extreme phenotypes, elevated disease risk, or earlier disease onset, and highlighted rare coding variants and regulatory elements that provide insight into underlying genetic risk factors, thereby complementing cvPRSs.

To date, PRS development has focused predominantly on cvPRSs derived from GWASs, owing to the widespread availability and cost-effectiveness of genotyping arrays and large-scale GWAS summary statistics^3,6–9^. However, cvPRSs capture only genetic variation tagged by genotyped and imputed variants and are often challenging to interpret due to pervasive linkage disequilibrium (LD) and highly polygenic architectures. In contrast, disease-associated rare variants typically exhibit larger effect sizes and are enriched in functionally important genomic regions^16–18^. Leveraging whole-exome and whole-genome sequencing enables the identification of rare coding and noncoding variants, including protein-truncating and damaging missense variants with large, biologically interpretable effects, as well as regulatory variants that may have modest individual effects but substantial cumulative impact through pathways and gene regulatory networks. Incorporating these signals into rvPRSs provides a complementary view of genetic risk across the full allele frequency spectrum that is largely inaccessible to array-based approaches.

Despite this promise, prior studies of rvPRSs have varied in construction methods, focused primarily on population-level metrics, and were limited to selected traits and diseases^20,21,23^. Here, by integrating functional annotations into variant grouping and aggregating variants into gene- and annotation-level burden scores, we established a principled and flexible framework for rvPRS construction that prioritizes variants with higher likelihood of functional impact and aligns with established rare-variant association testing frameworks^17,18,25^. Combined with machine learning models, this framework enables scalable integration of rare-variant signals and facilitates interpretation through feature importance analyses, highlighting genes, regulatory elements, and pathways that drive predictive performance.

Consistent with theoretical expectations and prior empirical observations^20,21,23^, rvPRSs contributed modestly to population-level metrics such as variance explained beyond cvPRSs, reflecting the low frequency of rare variants. However, their impact varied across traits and diseases and was strongly influenced by genetic architecture. For phenotypes driven by large-effect mutations in specific genes – for example, protein-truncating and damaging missense variants in *APOC3* for HDL cholesterol or *TTN* for heart failure – rvPRSs provided substantial predictive contributions, in some cases comparable to or exceeding those of cvPRSs.

Beyond average prediction metrics, rvPRS improved the identification of individuals at the extremes of risk. For many traits and diseases, rvPRSs identified distinct – and sometimes larger – sets of high-risk individuals compared with cvPRSs. These individuals were often carriers of rare variants with large functional effects in disease-relevant genes that were not captured by common-variant analyses. For noncoding rvPRSs, distinct individuals were sometimes enriched for regulatory elements also implicated by cvPRSs, suggesting convergence of rare- and common-variant signals on shared biological pathways. Together, these findings indicate that rvPRSs not only enhance risk stratification but also provide insight into underlying disease mechanisms. Survival analyses further demonstrated that rvPRSs provide additional stratification of disease trajectories beyond cvPRSs over extended follow-up, supporting their prospective relevance.

This improved discrimination at the tails of the risk distribution has important clinical implications, as individuals with extreme genetic burden may benefit most from targeted screening and preventive interventions. For several traits and diseases, individuals in the top 1% of the rvPRS distribution exhibited substantially elevated risk relative to the reminder of the sample, in some cases exceeding the effect observed for cvPRSs. Moreover, rvPRSs can implicate specific genes, pathways, and regulatory networks, providing biologically interpretable signals that may inform disease subtypes, therapeutic targets, and prognosis. Future work integrating longitudinal clinical data will be important for characterizing disease trajectories, treatment response, and survival outcomes among individuals identified by rvPRSs, thereby facilitating translation into precision medicine.

This study has several limitations. First, statistical power remains limited for rarer diseases. We anticipate that rvPRSs may play an even larger role in predicting and stratifying lower-prevalence diseases, which are more likely to be influenced by rare variants. Second, PRSs were developed and evaluated within the UK Biobank. Validation in independent cohorts is needed to assess generalizability, particularly given differences in variant sets and sample characteristics. Third, our analyses were restricted to individuals of predominantly European ancestry. Although rvPRS may be less sensitive to differences in LD patterns across populations and thus more portable than cvPRSs, their transferability to diverse ancestries warrants further investigation.

In summary, by leveraging whole-genome sequencing, functional annotations, and machine learning, we provide a comprehensive framework for constructing, evaluating, and interpreting rvPRSs. Our results demonstrate that rvPRSs complement cvPRSs, particularly for identifying individuals at the extremes of risk and for elucidating the genetic architecture of complex traits and diseases. As sequencing data become increasingly available in biobank-scale cohorts and clinical settings, integrating rare-variant information into polygenic prediction will be essential for developing models of genetic risk across the allele frequency spectrum and advancing precision medicine^4,80,81^.

## Methods

### UK Biobank

The UK Biobank (UKB) is a large population-based prospective cohort that enrolled approximately 500,000 participants aged 40-69 years at baseline between 2006 and 2010^82^. Ethical approval was obtained from the North West Multi-centre Research Ethics Committee. All participants provided informed consent. Extensive data were collected, including whole-genome sequencing data, questionnaire responses, physical measurements, and linked national health records, with more than 15 years of follow-up. The current study was conducted under UKB application 19542.

### Processing and quality control of whole-genome sequencing data

Whole-genome sequencing (WGS) data in the UKB were generated by deCODE Genetics using paired-end sequencing on Illumina NovaSeq 6000 instruments, with an average coverage of 32.5×^83,84^. Detailed sequencing procedures and initial quality control have been described previously^83^. Variants were called using the Illumina DRAGEN pipeline (v3.7.8) and aligned to the GRCh38 reference genome. In this study, we used the joint-called pVCF WGS data (field 24310) and performed additional quality control as described below.

Multiallelic variants were split into biallelic variants and normalized using bcftools^85^. We applied a series of variant-level filters, excluding SNPs and indels with any of the following: (i) missingness >10% across individuals; (ii) heterozygosity excess outside the range 0.5-1.5; (iii) missing alternative alleles; or (iv) allele frequencies outside the valid range 0-1. Only variants with “FILTER = PASS” were retained.

At the sample level, the cohort was restricted to unrelated individuals of White British ancestry (field ID 22006), defined using a kinship coefficient <0.0884. Hardy-Weinberg equilibrium was assessed after sample restriction, and variants with P <10^−30^ were excluded. We further excluded participants who withdrew consent or showed discrepancies between genetically-inferred and self-reported sex. Additional sample-level filters removed individuals with extreme transition/transversion (Ti/Tv) or heterozygous/homozygous (Het/Hom) ratios >8 standard deviations, call rate <95%, or outlying total variant counts.

After quality control, the dataset comprised 1,023,836,503 variant sites across 345,967 individuals, including 6,290,920 common variants (MAF ≥1%) and 1,017,545,583 rare variants (MAF <1%).

### Functional annotation of coding and noncoding rare variants

We defined 15 gene-centric functional categories following the annotation framework implemented in the STAARpipeline^17,18,25^.

For coding regions, rare variants within protein-coding genes were aggregated into seven categories: (i) predicted loss-of function (pLoF) variants, including stop-gain, stop-loss, frameshift, and splice-site variants; (ii) protein-truncating variants (PTVs); (iii) missense variants; (iv) disruptive missense variants; (v) pLoF + disruptive missense variants; (vi) PTVs + disruptive missense variants; and (vii) synonymous variants. The pLoF, missense, and synonymous categories were defined using GENCODE Variant Effect Predictor (VEP) annotations^86,87^. For missense variants, we defined the disruptive subset using MetaSVM^88^, a meta-analytic support vector machine that predicts the deleteriousness of missense mutations.

For noncoding regions, rare variants were aggregated into eight gene-centric regulatory categories: (i) promoter variants overlapping CAGE sites; (ii) promoter variants overlapping DHS sites; (iii) enhancer variants overlapping CAGE sites; (iv) enhancer variants overlapping DHS sites; (v) untranslated regions (UTRs); (vi) upstream regions; (vii) downstream regions; and (viii) noncoding RNA (ncRNA) genes. Promoter variants were defined as rare variants located within ±3 kilobase (kb) of transcription start sites. Enhancer variants were defined as rare variants located in GeneHancer-predicted enhancer regions^89–92^. Variants in UTRs, upstream, downstream, and ncRNA categories were defined using GENCODE VEP annotations^86,87^. The UTR category included both 5′ and 3′ UTR variants, and the ncRNA category included exonic and splice-region variants in noncoding RNA genes. Definitions for the first seven categories were obtained from Ensembl^93^, and ncRNA gene annotations were obtained from GENCODE^86,87^.

### Quantitative traits

We analyzed 31 quantitative traits, including anthropometric measurements and blood biomarkers (Supplementary Table 1). Each phenotype was first adjusted for common covariates, including age, sex, and top 10 genetic principal components (PCs), and was subsequently standardized using the mean and variance estimated in the training set.

### Disease endpoints

In total, we analyzed 464 disease endpoints^94^, each with at least 466 cases among participants with available WGS data (Supplementary Table 2). Disease endpoints were defined based on the earliest recorded instance of a three-character ICD-10 code, following the FinnGen clinical endpoint framework (Data Freeze 12). These codes classify diseases into 14 chapters, including infectious diseases, neoplasms, hematologic and immune disorders, endocrine and metabolic conditions, psychiatric disorders, nervous system diseases, ophthalmic conditions, circulatory diseases, respiratory diseases, gastrointestinal disorders, dermatologic conditions, musculoskeletal diseases, genitourinary disorders, and pregnancy-related outcomes. Disease data were extracted from hospital inpatient records (field IDs 41270 and 41280) obtained from UK Hospital Episode Statistics.

We restricted analyses to incident disease endpoints, defined as diagnoses occurring after the baseline assessment, corresponding to the date of blood sample collection. Follow-up began at baseline and continued until the earliest of first diagnosis of the endpoint, death, or the last available inpatient record (November 2023). Individuals with a diagnosis of the endpoint before baseline were excluded. Controls were defined as participants who remained free of the disease throughout follow-up. Disease data underwent quality control based on FinnGen guidelines, including sex- and age-specific criteria and predefined inclusion and exclusion thresholds.

### Gene-level burden scores and feature pre-filtering

For each gene and MAF threshold, variants within each coding or noncoding category defined above and with MAF below the threshold were aggregated into burden scores, yielding 15 features per gene (7 coding and 8 noncoding) across 18,445 genes. To obtain a tractable set of input features for downstream PRS modeling, we performed univariate pre-filtering by computing the Pearson correlation between each burden score and the phenotype, retaining features with *p* <0.001. The selected burden scores were then used as input features for subsequent PRS construction models.

### rvPRS construction models

We evaluated five approaches for rvPRS construction:

(i) Linear models. Linear regression was used for quantitative traits and balanced logistic regression, with inverse case/control sample-size weights, was used for disease endpoints.
(ii) Regularized linear models (GLMNET). Elastic net regularization was applied to both linear regression and balanced logistic regression. Hyperparameters, including the overall regularization strength and the relative contributions of LASSO and ridge penalties, were selected based on prediction accuracy in the validation set, using correlation *R* for quantitative traits and AUC for disease endpoints.
(iii) Multi-layer perceptron (MLP). We implemented a feedforward neural network with one hidden layer. Models were trained using mean square error (MSE) loss for quantitative traits and weighted cross-entropy (WCE) loss for disease endpoints. Hyperparameters, including gradient accumulation steps and width of the hidden layer, were tuned based on validation performance. Additional implementation details are provided in Supplementary Methods and Supplementary Table 18.
(iv) Light gradient boosting machines (LightGBM). LightGBM is a tree-based method that captures nonlinear effects and higher-order interactions without explicit feature engineering. Models were trained using MSE loss for quantitative traits and WCE loss for disease endpoints. Hyperparameters, including the number of leaves, learning rate, and minimum data of observations per leaf, were selected based on validation performance. Feature importance was assessed using SHapley Additive exPlanatory (SHAP) values^32,33^. Additional implementation details are provided in Supplementary Methods and Supplementary Table 19.
(v) Transformer-based rvPRS model (RVTrans). We implemented a Transformer-based^30,31^ rvPRS model incorporating a self-attention feature-ranking mechanism that represents burden scores as embeddings informed by their statistical association with the phenotype (Supplementary Figure 10). This approach groups functionally related features and prioritizes informative, potentially dispersed signals, improving predictive performance. Models were trained using HL-Gauss loss^95,96^ for quantitative traits and balanced cross-entropy loss for disease endpoints. Hyperparameters, including gradient accumulation steps, patch size, and model depth, were tuned based on validation performance. Feature importance was assessed using saliency maps^97^. Additional implementation details are provided in Supplementary Methods and Supplementary Table 20.

### Common-variant GWAS and cvPRS construction

We performed common-variant GWAS for each quantitative trait and disease endpoint using the training set and approximately 620,000 common variants (MAF >1%) from the HapMap3 panel that passed quality control in the WGS dataset (Supplementary Tables 1-2). All GWAS analyses adjusted for age, sex, and the top 10 genetic PCs. The resulting GWAS summary statistics were used as input to PRS-CS-auto^3,6^ for cvPRS construction, with linkage disequilibrium (LD) reference panels derived from the 1000 Genomes Project^98^ European samples. PRS-CS-auto infers all model parameters directly from data and therefore does not require hyperparameter tuning. Posterior effect size estimates were then applied to the testing set to construct cvPRSs.

### Population-level performance metrics for rvPRSs

We evaluated the predictive performance of rvPRSs using three complementary metrics:

(i) Prediction accuracy. We quantified prediction accuracy using the correlation *R*, defined as the square root of the incremental variance explained by the rvPRS beyond the cvPRS and covariates, including age, sex, and top 10 genetic PCs. For disease endpoints, this quantity was further transformed to the liability scale^99,100^.
(ii) Tail discrimination. We assessed discrimination at the extremes by estimating the odds ratio (OR) comparing individuals in the top 1% of the rvPRS distribution with the remainder of the sample, adjusting for the cvPRS and covariates, including age, sex, and top 10 genetic PCs. For quantitative traits, extreme phenotypes were defined as individuals in the top 1% of the phenotype distribution.
(iii) Integrated discrimination improvement (IDI)^101^. We evaluated whether adding the rvPRS to a model already including cvPRS and covariates improves case-control discrimination using IDI. IDI was defined as the change in discrimination slope, measured as the difference in average predicted probability between cases and controls, between models with and without the rvPRS. For quantitative traits, extreme phenotypes were defined as individuals in the top 1% of the phenotype distribution. Case probabilities were estimated using logistic regression.

### PRS integration

We linearly combined coding and noncoding rvPRSs to obtain a single combined rvPRS. Regression coefficients were estimated in the validation set using standardized coding and noncoding PRSs and were then applied to the testing set to compute the combined rvPRS. Similarly, the combined rvPRS and cvPRS were integrated using linear models to construct a unified PRS.

### Survival analysis

Associations between rvPRSs and time to disease onset were evaluated using Cox proportional hazards models, adjusting for age, sex, birth year, top 10 genetic PCs, and the cvPRS derived from PRS-CS. *P*-values were obtained from Wald tests of the rvPRS regression coefficients. Cox models were fitted using the *coxph* function from the *survival* package in R. Kaplan-Meier curves were generated for participants in the top 1%, middle 20%, and bottom 1% of the rvPRS distribution. Differences between strata were evaluated using log-rank tests, with survival curves estimated using the *survfit* function from the *survminer* package in R. Odds ratios were derived from logistic regression models comparing individuals in the top 1% of the rvPRS distribution with those in the middle 20% or bottom 1% strata. Corresponding *p*-values were obtained from Wald tests of the binary PRS-stratum coefficient, adjusting for age, sex, birth year, top 10 genetic PCs, and the cvPRS.

## Acknowledgements

This work was supported by Noncommunicable Chronic Diseases-National Science and Technology Major Project (2025ZD0546300) and the National Natural Science Foundation of China (Grant No. 82402379).

## Competing Interests

The authors declare no competing interests.

## Data Availability

Access to UK Biobank (UKB) data can be requested through the UK Biobank Access Management System: https://ams.ukbiobank.ac.uk/ams. UKB data used in this study were obtained under application 19542. *All of Us* WGS association summary statistics are available at https://allbyall.researchallofus.org.

## Code Availability

The following software and resources were used in this study:

STAAR: https://github.com/li-lab-genetics/STAAR

PRS-CS-auto: https://github.com/getian107/PRScs

PLINK2 (v2.00a3LM): https://www.cog-genomics.org/plink/2.0

Study-specific code: https://github.com/qin1114/Rare-variant-PRS/tree/main

Analyses were performed using Python 3.9.21, lightgbm 4.6.0, PyTorch 2.5.1, and CUDA 12.1.

## Notes

### Competing Interest Statement

The authors have declared no competing interest.

### Author Declarations

This study used UK Biobank data obtained under application 19542 and All of Us whole-genome sequencing association summary statistics that are publicly available.

## References

1. Chatterjee, N., Shi, J. & García-Closas, M. Developing and evaluating polygenic risk prediction models for stratified disease prevention. Nature Reviews Genetics 17, 392–406 (2016).

2. Khera, A.V. et al. Genome-wide polygenic scores for common diseases identify individuals with risk equivalent to monogenic mutations. Nature Genetics 50, 1219–1224 (2018).

3. Ruan, Y. et al. Improving polygenic prediction in ancestrally diverse populations. Nature Genetics 54, 573–580 (2022).

4. Torkamani, A., Wineinger, N.E. & Topol, E.J. The personal and clinical utility of polygenic risk scores. Nature Reviews Genetics 19, 581–590 (2018).

5. Kullo, I.J. Clinical use of polygenic risk scores: current status, barriers and future directions. Nature Reviews Genetics (2025).

6. Ge, T., Chen, C.-Y., Ni, Y., Feng, Y.-C.A. & Smoller, J.W. Polygenic prediction via Bayesian regression and continuous shrinkage priors. Nature Communications 10, 1776 (2019).

7. Privé, F., Arbel, J. & Vilhjálmsson, B.J. LDpred2: better, faster, stronger. Bioinformatics 36, 5424–5431 (2021).

8. Privé, F., Arbel, J., Aschard, H. & Vilhjálmsson, B.J. Identifying and correcting for misspecifications in GWAS summary statistics and polygenic scores. HGG Adv 3, 100136 (2022).

9. Privé, F., Albiñana, C., Arbel, J., Pasaniuc, B. & Vilhjálmsson, B.J. Inferring disease architecture and predictive ability with LDpred2-auto. Am J Hum Genet 110, 2042–2055 (2023).

10. Van Hout, C.V. et al. Exome sequencing and characterization of 49,960 individuals in the UK Biobank. Nature 586, 749–756 (2020).

11. Karczewski, K.J. et al. The mutational constraint spectrum quantified from variation in 141,456 humans. Nature 581, 434–443 (2020).

12. Weiner, D.J. et al. Polygenic architecture of rare coding variation across 394,783 exomes. Nature 614, 492–499 (2023).

13. Sun, B.B. et al. Genetic associations of protein-coding variants in human disease. Nature 603, 95–102 (2022).

14. Backman, J.D. et al. Exome sequencing and analysis of 454,787 UK Biobank participants. Nature 599, 628–634 (2021).

15. Wang, Q. et al. Rare variant contribution to human disease in 281,104 UK Biobank exomes. Nature 597, 527–532 (2021).

16. Mbatchou, J. et al. Computationally efficient whole-genome regression for quantitative and binary traits. Nature Genetics 53, 1097–1103 (2021).

17. Li, X. et al. Dynamic incorporation of multiple in silico functional annotations empowers rare variant association analysis of large whole-genome sequencing studies at scale. Nat Genet 52, 969–983 (2020).

18. Li, Z. et al. A framework for detecting noncoding rare-variant associations of large-scale whole-genome sequencing studies. Nat Methods 19, 1599–1611 (2022).

19. Demontis, D. et al. Rare genetic variants confer a high risk of ADHD and implicate neuronal biology. Nature 649, 909–917 (2026).

20. Chen, T., Li, X., Zhang, H., Mazumder, R. & Lin, X. STELLAR: A flexible ensemble learning framework integrating rare variants to enhance polygenic risk prediction. medRxiv, 2026.06.07.26355109 (2026).

21. Williams, J. et al. Integrating common and rare variants improves polygenic risk prediction across diverse populations. Nature Communications (2026).

22. Souaiaia, T. et al. Distinct genetic architecture in the tails of complex traits. Nature (2026).

23. Fiziev, P.P. et al. Rare penetrant mutations confer severe risk of common diseases. Science 380, eabo1131 (2023).

24. Clarke, B. et al. Integration of variant annotations using deep set networks boosts rare variant association testing. Nat Genet 56, 2271–2280 (2024).

25. Li, X. et al. Streamlining large-scale genomic data management: Insights from the UK Biobank whole-genome sequencing data. Cell Genom, 101009 (2025).

26. Dong, C. et al. Comparison and integration of deleteriousness prediction methods for nonsynonymous SNVs in whole exome sequencing studies. Hum Mol Genet 24, 2125–37 (2015).

27. Zou, H. & Hastie, T. Regularization and variable selection via the elastic net. Journal of the Royal Statistical Society Series B: Statistical Methodology 67, 301–320 (2005).

28. Duffy, Á. et al. Development of a human genetics-guided priority score for 19,365 genes and 399 drug indications. Nature Genetics 56, 51–59 (2024).

29. Deng, Y. et al. Atlas of the plasma proteome in health and disease in 53,026 adults. Cell 188(2024).

30. Dosovitskiy, A., et al. An image is worth 16x16 words: Transformers for image recognition at scale. arXiv preprint arXiv:2010.11929 (2020).

31. Vaswani, A. et al. Attention is all you need. Advances in neural information processing systems 30(2017).

32. Lundberg, S.M. et al. From local explanations to global understanding with explainable AI for trees. Nature Machine Intelligence 2, 56–67 (2020).

33. Lundberg, S.M. & Lee, S.-I. A unified approach to interpreting model predictions. Advances in neural information processing systems 30(2017).

34. Angel, G.d., et al. P027: Genetic landscape of ALPL and other genes impacting alkaline phosphatase variation in the UK Biobank. Genetics in Medicine Open 3, 102871 (2025).

35. Bick, A.G. et al. Genomic data in the All of Us Research Program. Nature 627, 340–346 (2024).

36. Lu, W. et al. Systematic common and rare variant association testing in 392,030 whole genomes in *All of Us*. medRxiv, 2026.05.08.26350964 (2026).

37. Pike, A.F., Kramer, N.I., Blaauboer, B.J., Seinen, W. & Brands, R. A novel hypothesis for an alkaline phosphatase ‘rescue’ mechanism in the hepatic acute phase immune response. Biochim Biophys Acta 1832, 2044–56 (2013).

38. Coni, S. et al. Blockade of EIF5A hypusination limits colorectal cancer growth by inhibiting MYC elongation. Cell Death & Disease 11, 1045 (2020).

39. Saif, M.W., Alexander, D. & Wicox, C.M. Serum Alkaline Phosphatase Level as a Prognostic Tool in Colorectal Cancer: A Study of 105 patients. J Appl Res 5, 88–95 (2005).

40. Frikke-Schmidt, R. Genetic variation in the ABCA1 gene, HDL cholesterol, and risk of ischemic heart disease in the general population. Atherosclerosis 208, 305–16 (2010).

41. Feitosa, M.F., Myers, R.H., Pankow, J.S., Province, M.A. & Borecki, I.B. LIPC variants in the promoter and intron 1 modify HDL-C levels in a sex-specific fashion. Atherosclerosis 204, 171–7 (2009).

42. Delgado-Lista, J. et al. Effects of variations in the APOA1/C3/A4/A5 gene cluster on different parameters of postprandial lipid metabolism in healthy young men. J Lipid Res 51, 63–73 (2010).

43. Sorkin, S.C. et al. APOA1 polymorphisms are associated with variations in serum triglyceride concentrations in hypercholesterolemic individuals. Clin Chem Lab Med 43, 1339–45 (2005).

44. Giammanco, A., Spina, R., Cefalù, A.B. & Averna, M. APOC-III: a Gatekeeper in Controlling Triglyceride Metabolism. Curr Atheroscler Rep 25, 67–76 (2023).

45. Garelnabi, M., Lor, K., Jin, J., Chai, F. & Santanam, N. The paradox of ApoA5 modulation of triglycerides: evidence from clinical and basic research. Clin Biochem 46, 12–9 (2013).

46. Semmler, L., Reiter-Brennan, C. & Klein, A. BRCA1 and Breast Cancer: a Review of the Underlying Mechanisms Resulting in the Tissue-Specific Tumorigenesis in Mutation Carriers. J Breast Cancer 22, 1–14 (2019).

47. Mehrgou, A. & Akouchekian, M. The importance of BRCA1 and BRCA2 genes mutations in breast cancer development. Med J Islam Repub Iran 30, 369 (2016).

48. Toss, A. et al. Management of PALB2-associated breast cancer: A literature review and case report. Clin Case Rep 11, e7747 (2023).

49. Man, X. et al. DNMT3A and DNMT3B in Breast Tumorigenesis and Potential Therapy. Front Cell Dev Biol 10, 916725 (2022).

50. Lim, J.Y. et al. DNMT3A haploinsufficiency causes dichotomous DNA methylation defects at enhancers in mature human immune cells. J Exp Med 218(2021).

51. Holmström, M.O. et al. Evidence of immune elimination, immuno-editing and immune escape in patients with hematological cancer. Cancer Immunol Immunother 69, 315–324 (2020).

52. Abu Aqel, Y., et al. Glucokinase (GCK) in diabetes: from molecular mechanisms to disease pathogenesis. Cell Mol Biol Lett 29, 120 (2024).

53. Wang, W. et al. Impact of polymorphisms on gene expression and splicing in response to exercise and diet-induced weight loss in human skeletal muscle tissues. Cell Genom 5, 100951 (2025).

54. Rigi-Ladiz, M.A. et al. DNA methylation and expression status of glutamate receptor genes in patients with oral squamous cell carcinoma. Meta Gene 20, 100555 (2019).

55. Xia, Z.N., Wang, X.Y., Cai, L.C., Jian, W.G. & Zhang, C. IGLL5 is correlated with tumor-infiltrating immune cells in clear cell renal cell carcinoma. FEBS Open Bio 11, 898–910 (2021).

56. Zhang, J.X. et al. Association of tricellulin expression with poor colorectal cancer prognosis and metastasis. Oncol Rep 44, 2174–2184 (2020).

57. Naka, K. et al. The lysophospholipase D enzyme Gdpd3 is required to maintain chronic myelogenous leukaemia stem cells. Nature Communications 11, 4681 (2020).

58. Hassan, M. & Wagdy, K. ASGR1 - a new target for lowering non-HDL cholesterol. Glob Cardiol Sci Pract 2016, e201614 (2016).

59. Basu, D. & Goldberg, I.J. Regulation of lipoprotein lipase-mediated lipolysis of triglycerides. Curr Opin Lipidol 31, 154–160 (2020).

60. Calandra, S., Priore Oliva, C., Tarugi, P. & Bertolini, S. APOA5 and triglyceride metabolism, lesson from human APOA5 deficiency. Curr Opin Lipidol 17, 122–7 (2006).

61. Zvintzou, E. et al. Pleiotropic effects of apolipoprotein C3 on HDL functionality and adipose tissue metabolic activity. J Lipid Res 58, 1869–1883 (2017).

62. Standl, M. et al. FADS1 FADS2 gene cluster, PUFA intake and blood lipids in children: results from the GINIplus and LISAplus studies. PLoS One 7, e37780 (2012).

63. Fujita, Y. et al. Hypercholesterolemia associated with splice-junction variation of inter-α-trypsin inhibitor heavy chain 4 (ITIH4) gene. Journal of Human Genetics 49, 24–28 (2004).

64. Xie, C. et al. BRCA2 gene mutation in cancer. Medicine (Baltimore) 101, e31705 (2022).

65. Lee, J.-H. Targeting the ATM pathway in cancer: Opportunities, challenges and personalized therapeutic strategies. Cancer Treatment Reviews 129, 102808 (2024).

66. Jiang, S. Tet2 at the interface between cancer and immunity. Communications Biology 3, 667 (2020).

67. Mobet, Y. et al. AKAP8 promotes ovarian cancer progression and antagonizes PARP inhibitor sensitivity through regulating hnRNPUL1 transcription. iScience 27, 109744 (2024).

68. Wang, X., Qin, D. & Liu, L. AKAP8 interacting with DDX5 to regulate R-loop balance involves in lung carcinoma cell growth. bioRxiv, 2023.11.28.568985 (2023).

69. Fujimoto, J., Aoki, I., Toyoki, H., Khatun, S. & Tamaya, T. Clinical implications of expression of ETS-1 related to angiogenesis in uterine cervical cancers. Ann Oncol 13, 1598–604 (2002).

70. Wang, S. et al. ETS-1 in tumor immunology: implications for novel anti-cancer strategies. Front Immunol 16, 1526368 (2025).

71. Fullenkamp, D.E. Regulation of TTN as a mechanism of and treatment for heart failure. J Clin Invest 135(2025).

72. Wang, X. et al. Cardiac disruption of SDHAF4-mediated mitochondrial complex II assembly promotes dilated cardiomyopathy. Nature Communications 13, 3947 (2022).

73. Shu, H. et al. The role of CD36 in cardiovascular disease. Cardiovasc Res 118, 115–129 (2022).

74. Shi, M. & Guo, X. Abstract 4354797: The Role and Mechanism of Receptor-like Protein LRTM1 in the Negative Regulation of Cardiomyocyte Regeneration. Circulation 152, A4354797–A4354797 (2025).

75. Vattathil, S.M. et al. A Genetic Study of Cerebral Atherosclerosis Reveals Novel Associations with NTNG1 and CNOT3. Genes (Basel) 12(2021).

76. Tang, Y.L., Dong, X.Y., Zeng, Z.G. & Feng, Z. Gene expression-based analysis identified NTNG1 and HGF as biomarkers for diabetic kidney disease. Medicine (Baltimore) 99, e18596 (2020).

77. Roider, E.M. & Fisher, D.E. The impact of MITF on melanoma development: news from bench and bedside. J Invest Dermatol 134, 16–17 (2014).

78. Masaki, S. et al. Myelodysplastic Syndrome-Associated SRSF2 Mutations Cause Splicing Changes by Altering Binding Motif Sequences. Front Genet 10, 338 (2019).

79. Moran-Crusio, K. et al. Tet2 loss leads to increased hematopoietic stem cell self-renewal and myeloid transformation. Cancer Cell 20, 11–24 (2011).

80. Zeng, J. & Visscher, P.M. Harnessing functional annotation to improve the accuracy and transferability of polygenic scores. Nature Reviews Genetics 26, 805–806 (2025).

81. Wainschtein, P. et al. Estimation and mapping of the missing heritability of human phenotypes. Nature 649, 1219–1227 (2026).

82. Sudlow, C. et al. UK biobank: an open access resource for identifying the causes of a wide range of complex diseases of middle and old age. PLoS Med 12, e1001779 (2015).

83. Halldorsson, B.V. et al. The sequences of 150,119 genomes in the UK Biobank. Nature 607, 732–740 (2022).

84. Li, S., Carss, K., Halldórsson, B. & Cortes, A. Whole-genome sequencing of half-a-million UK Biobank participants, (2023).

85. Danecek, P. et al. Twelve years of SAMtools and BCFtools. Gigascience 10, (2021).

86. Harrow, J. et al. GENCODE: the reference human genome annotation for The ENCODE Project. Genome Res 22, 1760–74 (2012).

87. Frankish, A. et al. GENCODE reference annotation for the human and mouse genomes. Nucleic Acids Res 47, D766–d773 (2019).

88. Dong, C. et al. Comparison and integration of deleteriousness prediction methods for nonsynonymous SNVs in whole exome sequencing studies. Human Molecular Genetics 24, 2125–2137 (2014).

89. Dunham, I. et al. An integrated encyclopedia of DNA elements in the human genome. Nature 489, 57–74 (2012).

90. Andersson, R. et al. An atlas of active enhancers across human cell types and tissues. Nature 507, 455–461 (2014).

91. Forrest, A.R.R. et al. A promoter-level mammalian expression atlas. Nature 507, 462–470 (2014).

92. Fishilevich, S. et al. GeneHancer: genome-wide integration of enhancers and target genes in GeneCards. Database (Oxford) 2017(2017).

93. Kinsella, R.J. et al. Ensembl BioMarts: a hub for data retrieval across taxonomic space. Database (Oxford) 2011, bar030 (2011).

94. Deng, Y.-T. et al. Atlas of the plasma proteome in health and disease in 53,026 adults. Cell 188, 253–271.e7 (2025).

95. Imani, E. & White, M. Improving regression performance with distributional losses. in International conference on machine learning 2157–2166 (PMLR, 2018).

96. Peng, H., Gong, W., Beckmann, C.F., Vedaldi, A. & Smith, S.M. Accurate brain age prediction with lightweight deep neural networks. Medical Image Analysis 68, 101871 (2021).

97. Simonyan, K., Vedaldi, A. & Zisserman, A. Deep inside convolutional networks: Visualising image classification models and saliency maps. arXiv preprint arXiv:1312.6034 (2013).

98. Auton, A. et al. A global reference for human genetic variation. Nature 526, 68–74 (2015).

99. Dempster, E.R. & Lerner, I.M. Heritability of Threshold Characters. Genetics 35, 212–236 (1950).

100. Lee, S.H., Goddard, M.E., Wray, N.R. & Visscher, P.M. A better coefficient of determination for genetic profile analysis. Genet Epidemiol 36, 214–24 (2012).

101. Pencina, M.J., D’Agostino, R.B., Sr., D’Agostino, R.B., Jr. & Vasan, R.S. Evaluating the added predictive ability of a new marker: from area under the ROC curve to reclassification and beyond. Stat Med 27, 157–72; discussion 207–12 (2008).

