## Supplementary Information for "Rare-variant risk scores complement common-variant polygenic scores for disease risk prediction and stratification"

**Supplementary Methods**

**Multi-Layer Perceptron.** We trained a multi-layer perceptron (MLP) with one hidden layer for 30 epochs with a fixed batch size of 64. Gradient accumulation was used to simulate a larger effective batch size. Model training was supervised using mean squared error (MSE) loss for quantitative traits and weighted cross-entropy (WCE) loss for disease endpoints. Hyperparameters, including gradient accumulation steps and hidden-layer width, were selected by grid search (Supplementary Table 18). We used the AdamW[1] optimizer with a peak learning rate of 0.0001, weight decay of 0.01, and beta parameters of 0.9 and 0.999. A cosine annealing scheduler with warm-up was used to adjust the learning rate throughout training. Gradient clipping with a maximum norm of 1 was applied to prevent gradient explosion. Final model selection was based on validation-set performance, using the correlation $R$ for quantitative traits and AUC for disease endpoints. All models were trained on a single NVIDIA A100 GPU using PyTorch 2.5.1 and CUDA 12.1.

**Tree-based nonlinear approach: LightGBM.** LightGBM is a gradient boosting framework based on decision trees that captures complex interactions and higher-order nonlinearities without requiring explicit feature engineering. LightGBM constructs an ensemble of decision trees sequentially, with each tree trained to minimize the residual errors of the previous ensemble, thereby iteratively refining predictive performance. To improve computational efficiency and scalability, LightGBM uses a histogram-based algorithm for split finding and a leaf-wise tree growth strategy with depth constraints, enabling the model to handle large-scale, high-dimensional genomic data. Importance of predictors in LightGBM was assessed using SHapley Additive exPlanatory (SHAP) values[2,3]. Hyperparameters, including the number of leaves, learning rate, and minimum number of observations per leaf, were selected by grid search (Supplementary Table 19). Models were trained using MSE loss for quantitative traits and WCE loss for disease endpoints for up to 1,000 boosting rounds, with early stopping applied if validation performance did not improve over 20 consecutive rounds. Final model selection was based on validation-set performance, using the correlation $R$ for quantitative traits and AUC for disease endpoints.

**Transformer-based rvPRS model: RVTrans.** A Transformer is a deep learning architecture that uses self-attention mechanisms to process input data represented as a sequence of embeddings (Supplementary Figure 10). For a given phenotype, the input to the Transformer model was a matrix $\boldsymbol{X} \in \mathbb{R}^{B \times M}$, where $B$ denotes batch size and $M$ denotes the number of features. Let $\boldsymbol{b}_{g_c}$ denote the vector of burden scores across individuals in the batch for the $c$-th functional annotation of gene $g$. The input matrix was constructed as

$$\boldsymbol{X} = \mathrm{sort}\left(\left\{\boldsymbol{b}_{g_c} \mid g_c \in \boldsymbol{S}\right\}\right),$$

where $\boldsymbol{S}$ denotes the set of phenotype-specific gene-annotation masks. The columns of $\boldsymbol{X}$ were sorted according to the strength of their association with the phenotype, based on Pearson correlation *p*-values computed in the training set.

For disease endpoints, the input matrix $\boldsymbol{X}$ was divided into nonoverlapping patches of fixed size $L$ and projected into a 512-dimensional latent space using a one-dimensional convolution operation:

$$\boldsymbol{Z} = \text{Conv1D}(\boldsymbol{X}),$$

where $\boldsymbol{Z} \in \mathbb{R}^{B\times(M/L)\times 512}$. Here, the Conv1D layer used kernel size and stride equal to $L$, acting as a patch-wise linear projection.

For quantitative traits, we first encoded burden scores in $\boldsymbol{X}$ as the sequence of learnable embeddings, analogous to word embeddings in natural language processing[4]:

$$\boldsymbol{E} = \boldsymbol{W}_{\text{embed}}[\boldsymbol{X}],$$

where $\boldsymbol{W}_{\text{embed}} \in \mathbb{R}^{V\times 512}$ represents the embedding matrix, with $V$ corresponding to the number of possible encoded burden values. The embedded sequence was then partitioned into patches and projected to the same latent dimension:

$$\boldsymbol{Z}^{(0)} = \text{Reshape}(\boldsymbol{E}), \qquad \boldsymbol{Z}^{(1)} = \text{Linear}\left(\boldsymbol{Z}^{(0)}\right).$$

The resulting dimensions were $\boldsymbol{E} \in \mathbb{R}^{B\times M\times 512}$, $\boldsymbol{Z}^{(0)} \in \mathbb{R}^{B\times(M/L)\times(L\times 512)}$, and $\boldsymbol{Z}^{(1)} \in \mathbb{R}^{B\times(M/L)\times 512}$.

The feature embeddings $\boldsymbol{Z}^{(1)}$ were then fed into a standard Vision Transformer (ViT)[5]. A learnable classification token ([CLS]) was prepended to the input sequence and served as a global representation for phenotype prediction. The sequence of embeddings was processed through multiple Transformer layers consisting of multi-head self-attention and feed-forward networks. Let $\boldsymbol{h}_i$ denote the final [CLS] representation for individual $i$. This representation was used to predict the phenotype, and the resulting prediction was treated as the individual-level rvPRS for downstream analysis.

<u>Training objective</u>: For quantitative traits, we used the HL-Gauss loss[6,7], a distributional regression objective that converts continuous targets into soft-label distributions, to mitigate overfitting and enhance representation

learning. For individual $i$, the loss minimizes the Kullback-Leibler (KL) divergence between the soft-label distribution and the model-predicted distribution, equivalently minimizing cross-entropy up to a constant:

$$L_{\text{quant},i} = -\sum_{k=1}^{K} p_{i,k} \log \hat{p}_{i,k},$$

where $p_{i,k}$ represents the probability mass of the soft label for individual $i$ in the $k$-th bin, and $\hat{p}_{i,k} = f_k(\boldsymbol{h}_i)$ denotes the model-predicted probability for that bin. Each phenotype was adjusted for covariates, including age, sex, and top 10 genetic principal components (PCs), and normalized using the training set.

The final prediction was calculated as the expected value under the model-predicted bin probabilities:

$$\hat{y}_i = \sum_{k=1}^{K} \hat{p}_{i,k} t_k,$$

where $t_k$ denotes the representative trait value, such as the bin center, for the $k$-th bin.

Suppose the target trait has support $[a, b]$, uniformly partitioned into $K$ bins with width $w_k$. For a truncated Gaussian soft-label distribution, $p_{i,k}$ was calculated as:

$$p_{i,k} = \frac{1}{C_i}\left(\operatorname{erf}\left(\frac{l_k + w_k - \mu_i}{\sqrt{2}\sigma}\right) - \operatorname{erf}\left(\frac{l_k - \mu_i}{\sqrt{2}\sigma}\right)\right),$$

with normalization constant

$$C_i = \operatorname{erf}\left(\frac{b - \mu_i}{\sqrt{2}\sigma}\right) - \operatorname{erf}\left(\frac{a - \mu_i}{\sqrt{2}\sigma}\right),$$

where $l_k$ is the left boundary of the $k$-th bin, $\mu_i$ is the normalized target value for individual $i$, $\sigma = 2$, and $K = 100$.

For disease endpoints, we used a balanced cross-entropy loss, with class weights inversely proportional to class frequencies, to mitigate class imbalance. For individual $i$, the predicted disease probability was defined as $q_i =$

$\sigma(f(\boldsymbol{h}_i))$, where $f(\cdot)$ is the predictive model and $\sigma(\cdot)$ is the sigmoid function. The balanced cross-entropy loss was defined as:

$$L_{\text{disease},i} = -[\omega_1 y_i \log(q_i) + \omega_0(1 - y_i)\log(1 - q_i)],$$

where $y_i$ is the binary disease label. The class weights $\omega_1 = N/2N_1$ and $\omega_0 = N/2N_0$ were calculated based on the number of case ($N_1$) and control ($N_0$) samples in the training set, with $N = N_1 + N_0$.

Feature importance and model training: Predictor importance in RVTrans was assessed using saliency maps[8], which quantify the gradient of the loss function with respect to each input feature. These gradients provide importance scores that reflect how small perturbations in the input would affect the model output. For quantitative traits, because the embedding lookup prevents direct gradient calculation with respect to the original input features, we first computed gradients with respect to the embedding vectors. The saliency score for each original input feature was then derived by averaging gradient magnitudes across all dimensions of its corresponding embedding vector.

Hyperparameter selection and implementation: The model was trained for 30 epochs with a fixed batch size of 64, using gradient accumulation to simulate a larger effective batch size. Gradient accumulation steps, patch size, and model depth were selected by grid search (Supplementary Table 20). We used the AdamW[1] optimizer with a peak learning rate of 0.0001, weight decay of 0.01, and beta parameters of 0.9 and 0.999. A cosine annealing scheduler with warm-up was used to adjust the learning rate throughout training. Gradient clipping with a maximum norm of 1 was applied to prevent gradient explosion. Final model selection was based on validation-set performance, using the correlation *R* for quantitative traits and AUC for disease endpoints. All models were trained on a single NVIDIA A100 GPU using PyTorch 2.5.1 and CUDA 12.1.

**References**


1. Loshchilov, I. & Hutter, F. Decoupled weight decay regularization. *arXiv preprint arXiv:1711.05101* (2017).
2. Lundberg, S.M. & Lee, S.-I. A unified approach to interpreting model predictions. in *Proceedings of the 31st International Conference on Neural Information Processing Systems* 4768–4777 (Curran Associates Inc., Long Beach, California, USA, 2017).
3. Lundberg, S.M. *et al.* From local explanations to global understanding with explainable AI for trees. *Nature Machine Intelligence* **2**, 56–67 (2020).

4. Mikolov, T., Sutskever, I., Chen, K., Corrado, G. & Dean, J. Distributed representations of words and phrases and their compositionality. in *Proceedings of the 27th International Conference on Neural Information Processing Systems - Volume 2* Vol. 2 3111–3119 (Curran Associates Inc., Lake Tahoe, Nevada, 2013).
5. Dosovitskiy, A. *et al.* An Image is Worth 16x16 Words: Transformers for Image Recognition at Scale. *ArXiv* abs/2010.11929 (2020).
6. Imani, E. & White, M. Improving regression performance with distributional losses. in *International conference on machine learning* 2157–2166 (PMLR, 2018).
7. Peng, H., Gong, W., Beckmann, C.F., Vedaldi, A. & Smith, S.M. Accurate brain age prediction with lightweight deep neural networks. *Medical Image Analysis* **68**, 101871 (2021).
8. Simonyan, K., Vedaldi, A. & Zisserman, A. Deep inside convolutional networks: Visualising image classification models and saliency maps. *arXiv preprint arXiv:1312.6034* (2013).

### Supplementary Figures

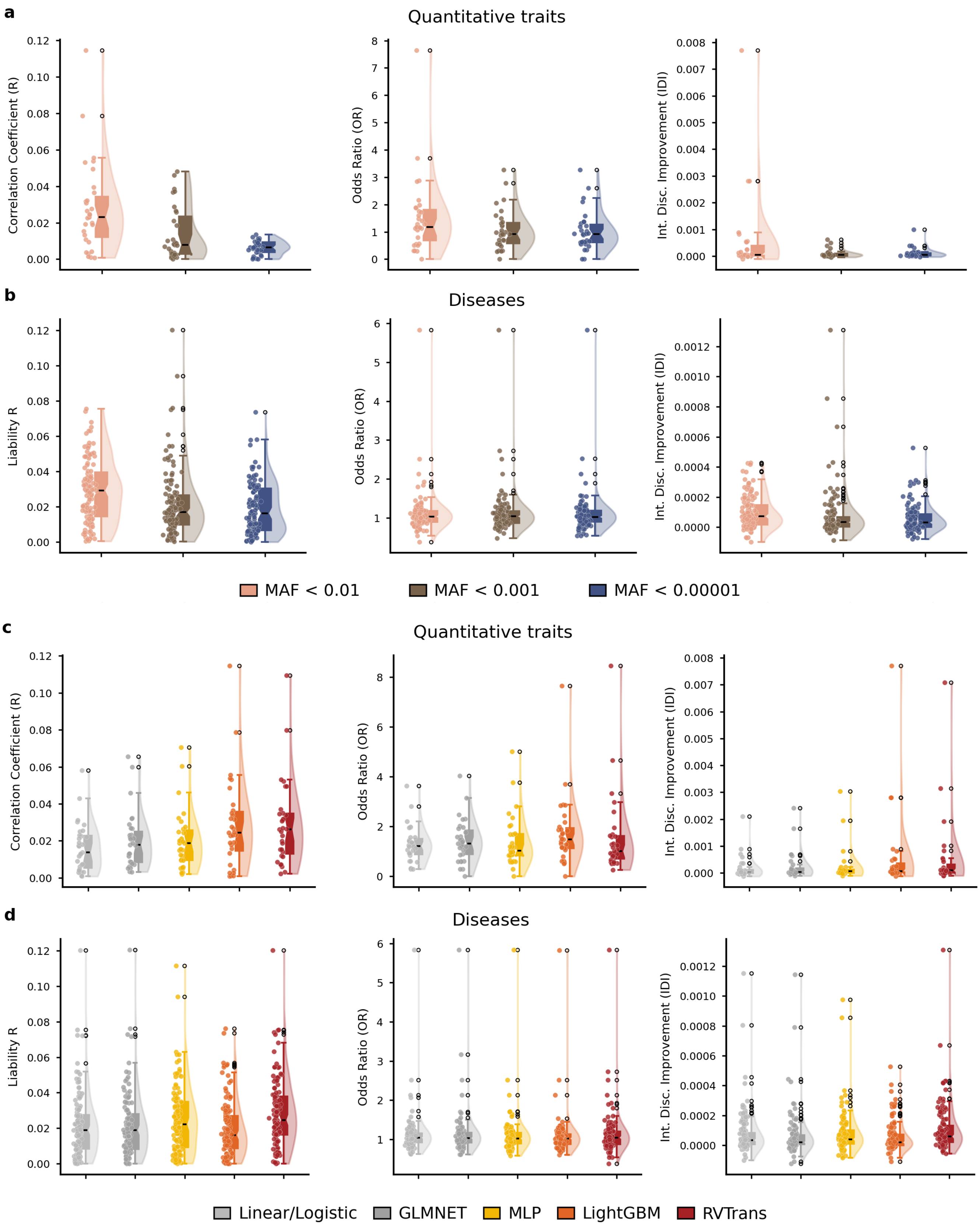

**Supplementary Figure 1: Population-level predictive performance of noncoding rare-variant polygenic risk scores across complex traits and diseases. a,** Incremental predictive performance of rvPRSs beyond cvPRSs for 31 quantitative traits, evaluated using three metrics: (i) the correlation *R* between predicted and observed phenotypes, (ii) the odds ratio (OR) comparing individuals in the top 1% of the rvPRS distribution with the remainder of the sample, and (iii) the integrated discrimination improvement (IDI), across three different minor allele frequency (MAF) cutoffs. **b,** Incremental predictive performance of rvPRSs beyond cvPRSs for 119 diseases with prevalence >5%, evaluated across three MAF cutoffs. **c,** Incremental predictive performance of rvPRSs beyond cvPRSs for 31 quantitative traits across five prediction models. **d,** Incremental predictive performance of rvPRSs beyond cvPRSs for 119 diseases with prevalence >5% across five prediction models. Across all panels, each data point represents the optimal result for a given trait or disease, selected across prediction models (for panels **a**-**b**), and across MAF cutoffs (for panels **c**-**d**). The center line of each box plot denotes the median, and box limits indicate the interquartile range.

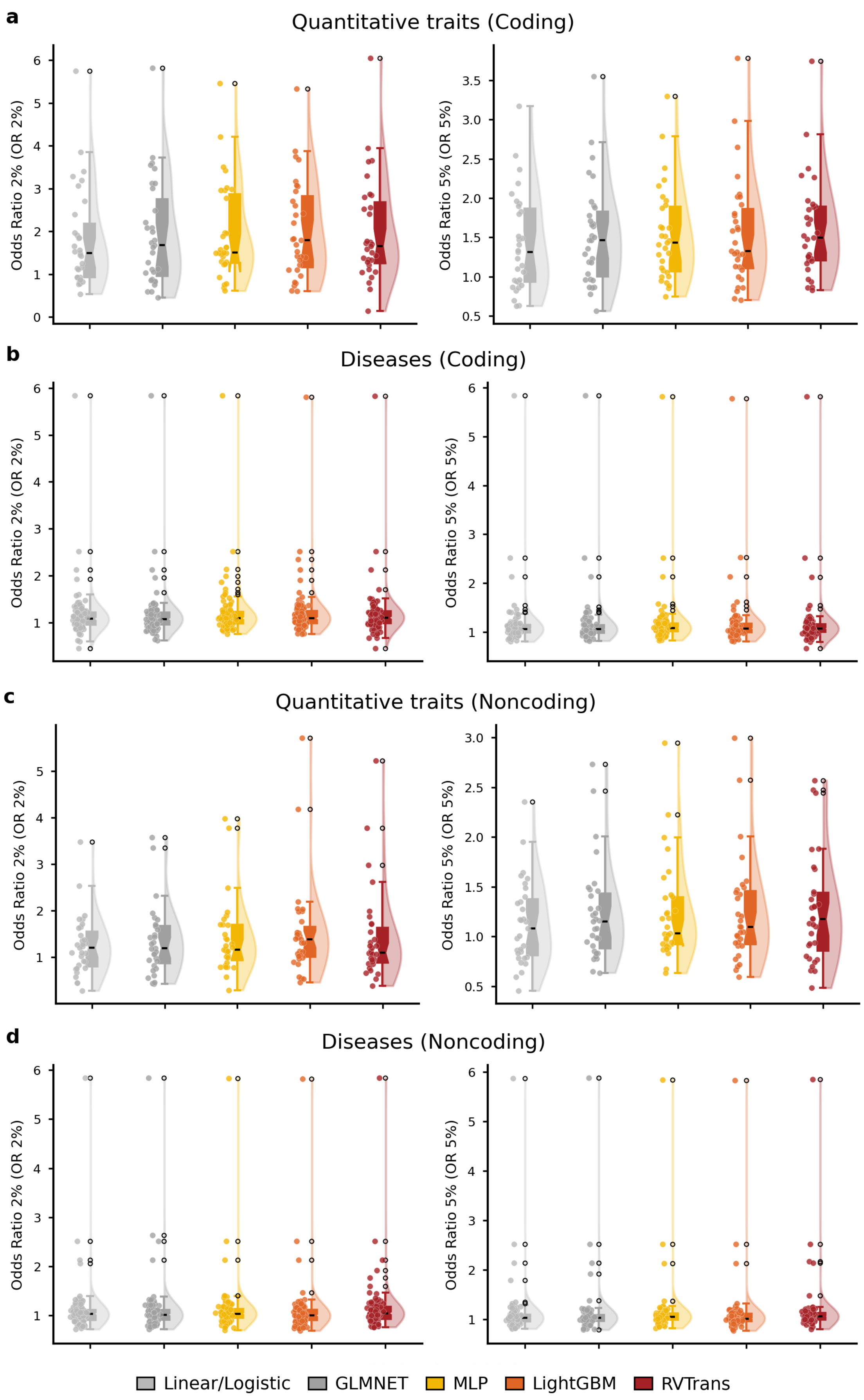
a
Quantitative traits (Coding)
Odds Ratio 2% (OR 2%)
Odds Ratio 5% (OR 5%)
b
Diseases (Coding)
Odds Ratio 2% (OR 2%)
Odds Ratio 5% (OR 5%)
c
Quantitative traits (Noncoding)
Odds Ratio 2% (OR 2%)
Odds Ratio 5% (OR 5%)
d
Diseases (Noncoding)
Odds Ratio 2% (OR 2%)
Odds Ratio 5% (OR 5%)
Linear/Logistic
GLMNET
MLP
LightGBM
RVTrans

**Supplementary Figure 2: Sensitivity analyses using alternative rare-variant polygenic risk score thresholds across complex traits and diseases. a,** Odds ratios comparing individuals in the top 2% or top 5% of the coding rvPRS distribution with the remainder of the sample for 31 quantitative traits across five prediction models. **b,** Odds ratios comparing individuals in the top 2% or top 5% of the coding rvPRS distribution with the remainder of the sample for 119 diseases with prevalence >5% across five prediction models. **c,** Odds ratios comparing individuals in the top 2% or top 5% of the noncoding rvPRS distribution with the remainder of the sample for 31 quantitative traits across five prediction models. **d,** Odds ratios comparing individuals in the top 2% or top 5% of the noncoding rvPRS distribution with the remainder of the sample for 119 diseases with prevalence >5% across five prediction models. Across all panels, each data point represents the optimal result for a given trait or disease, selected across MAF cutoffs. The center line of each box plot denotes the median, and box limits indicate the interquartile range.

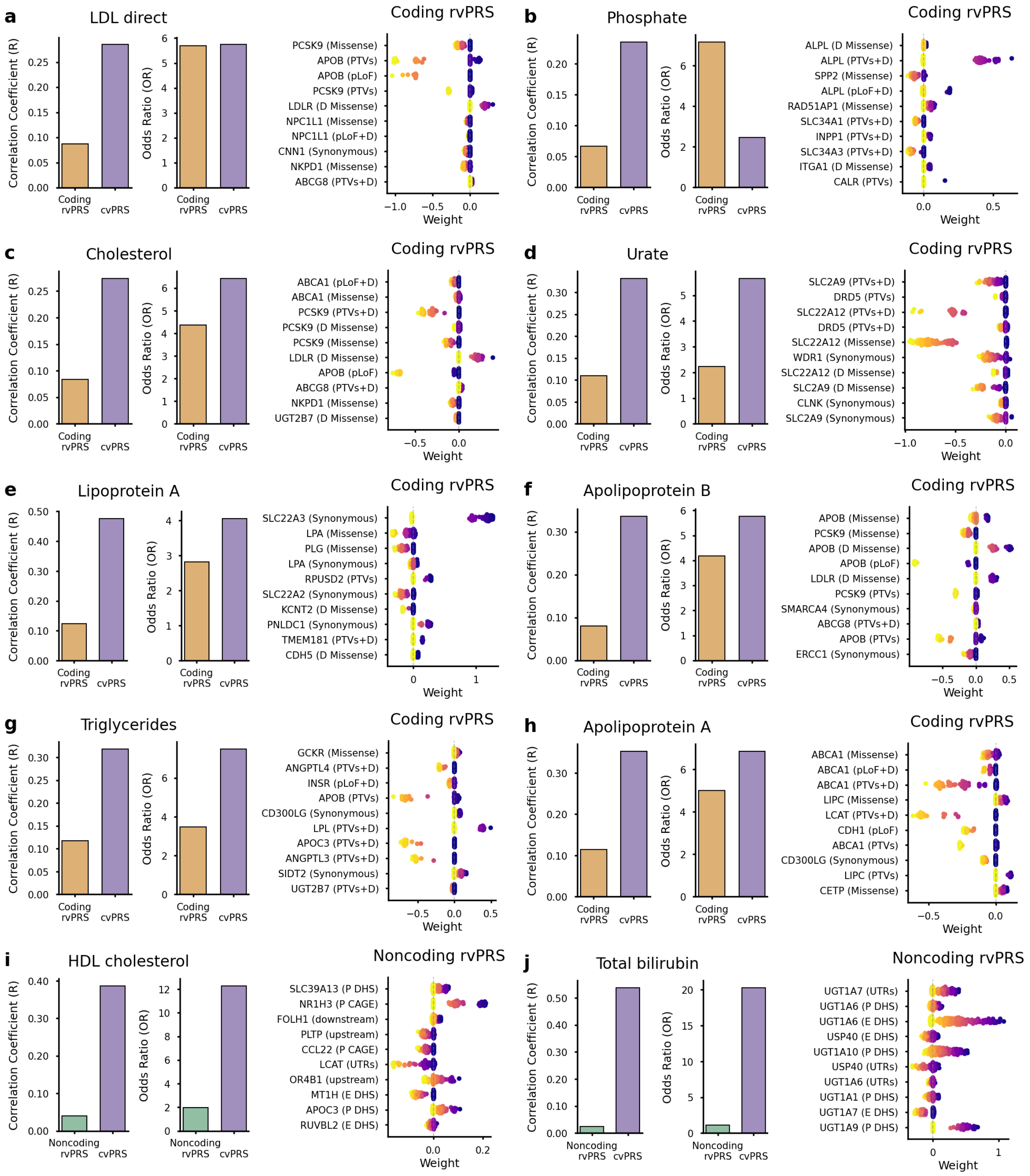


**Supplementary Figure 3: Additional examples of large-effect genes driving predictive performance of rare-variant polygenic risk scores for selected quantitative traits.** For each trait, the left panel shows prediction accuracy and risk discrimination for rvPRSs and cvPRSs, with discrimination quantified by the odds ratio comparing individuals in the top 1% of the corresponding PRS distribution with the reminder of the sample. The right panel shows the genes and functional annotations with the strongest contributions to rvPRS prediction, based on SHapley Additive exPlanations (SHAP) feature importance analysis implemented in the LightGBM models.

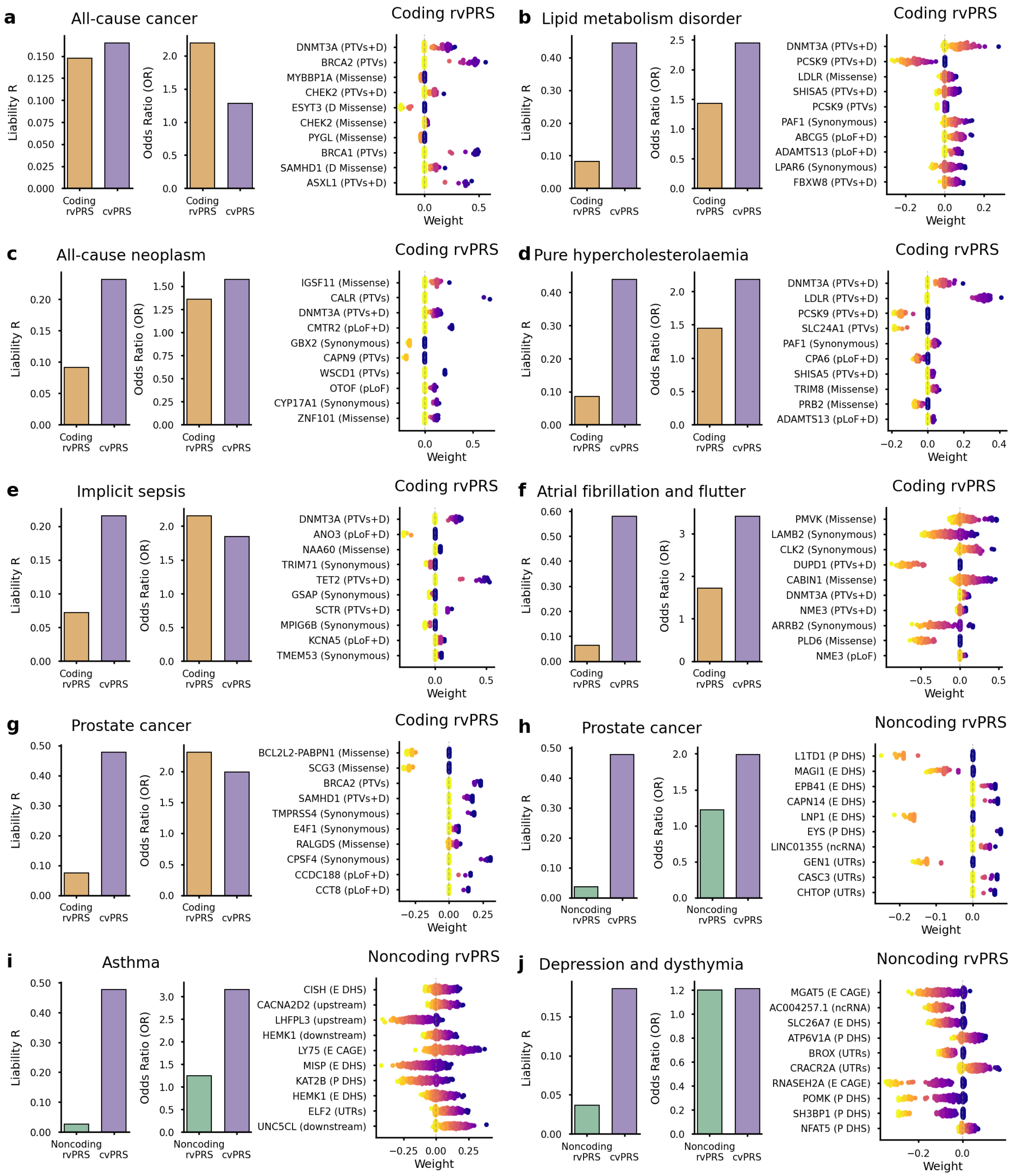


**Supplementary Figure 4: Additional examples of large-effect genes driving predictive performance of rare-variant polygenic risk scores for selected disease endpoints.** For each disease, the left panel shows prediction accuracy and risk discrimination for rvPRSs and cvPRSs, with discrimination quantified by the odds ratio comparing individuals in the top 1% of the corresponding PRS distribution with the reminder of the sample. The right panel shows the genes and functional annotations with the strongest contributions to rvPRS prediction, based on SHapley Additive exPlanations (SHAP) feature importance analysis implemented in the LightGBM models.

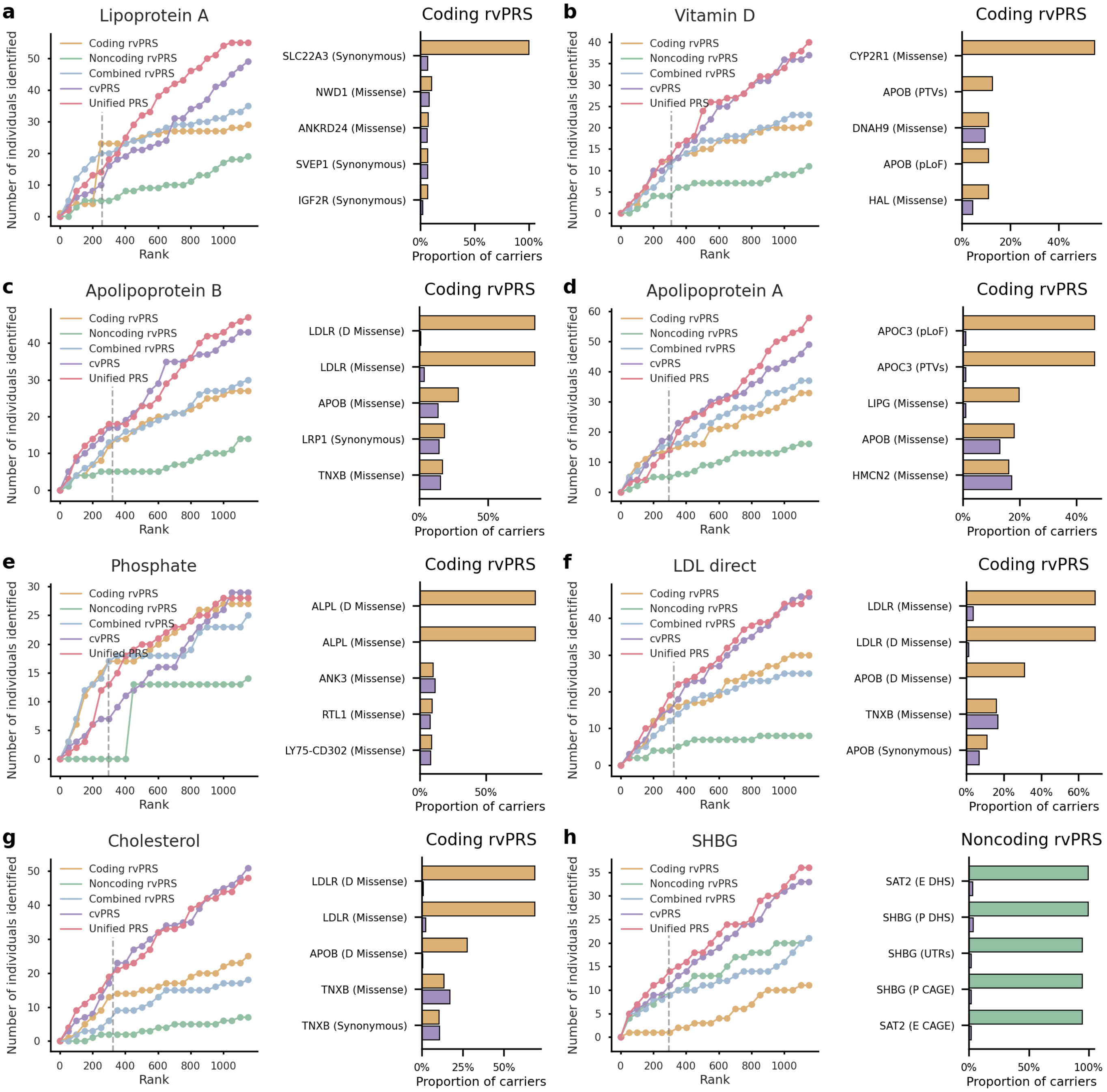


**Supplementary Figure 5: Additional examples of individuals with extreme phenotypes identified using rare-variant, common-variant, and integrated polygenic risk scores for selected quantitative traits.** For each trait, the left panel shows the number of individuals whose phenotype falls within the top 1% of the testing sample as a function of PRS rank for the coding rvPRS, noncoding rvPRS, combined rvPRS, cvPRS, and unified PRS integrating rvPRSs and cvPRSs. The vertical dashed gray line indicates the top 1% threshold of the PRS distribution. The right panel shows genes and functional annotations enriched among individuals identified by the coding rvPRS, noncoding rvPRS, and cvPRS.

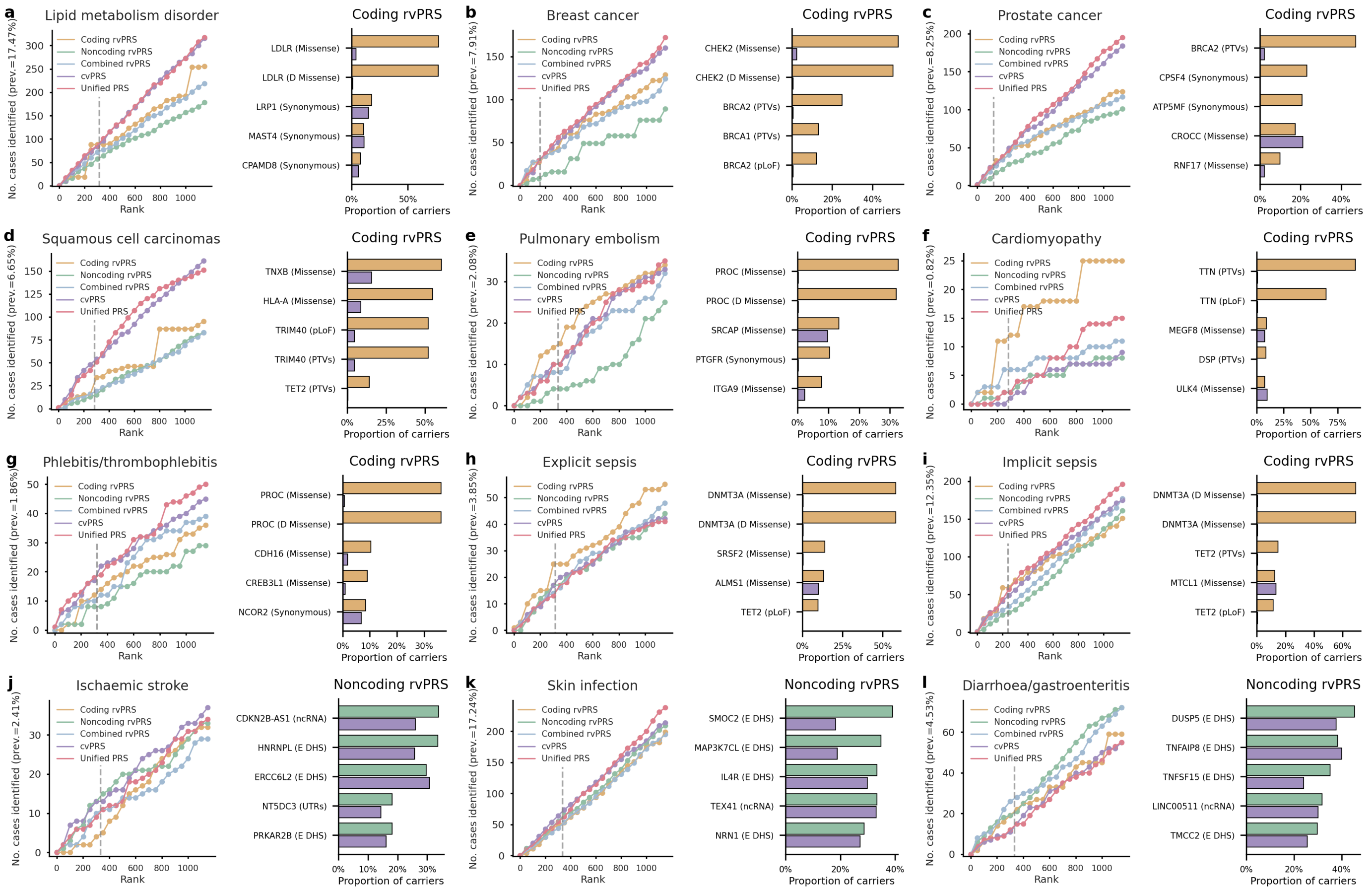


**Supplementary Figure 6: Additional examples of high-risk individuals identified using rare-variant, common-variant, and integrated polygenic risk scores for selected disease endpoints.** For each disease, the left panel shows the number of cases as a function of PRS rank for the coding rvPRS, noncoding rvPRS, combined rvPRS, cvPRS, and unified PRS integrating rvPRSs and cvPRSs. The vertical dashed gray line indicates the top 1% threshold of the PRS distribution. The right panel shows genes and functional annotations enriched among individuals identified by the coding rvPRS, noncoding rvPRS, and cvPRS.

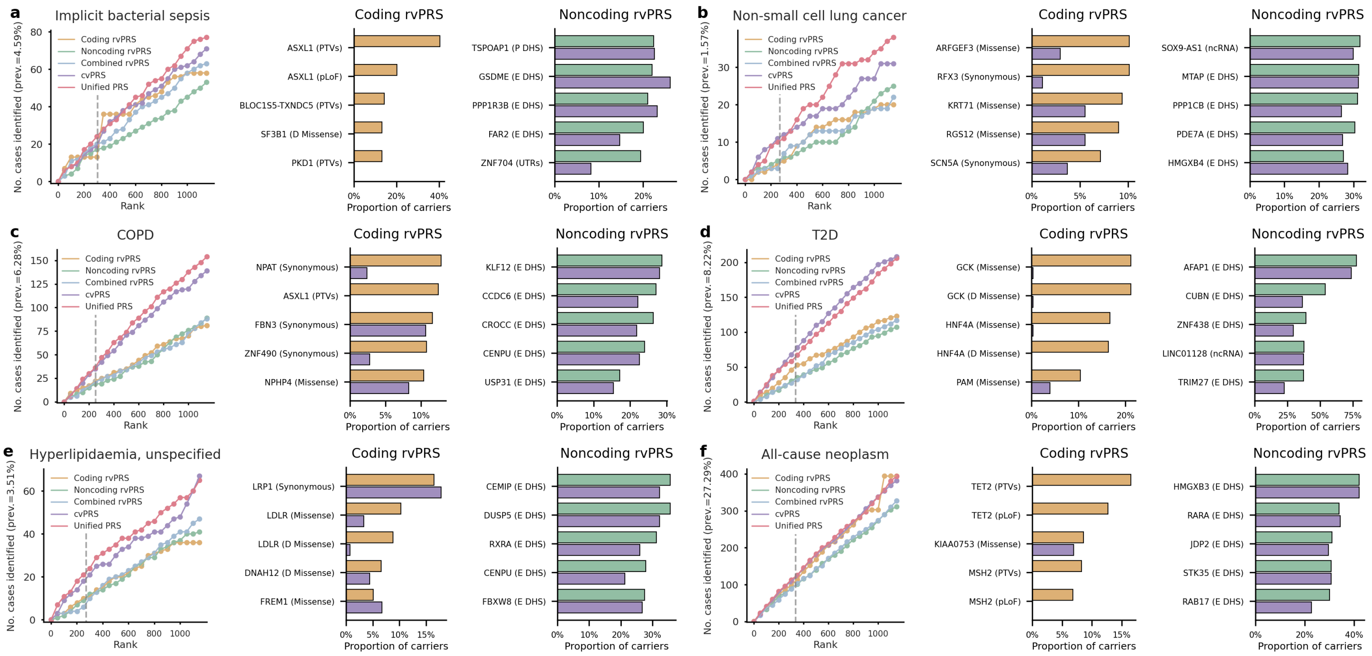


**Supplementary Figure 7: Additional examples of high-risk individuals identified using rare-variant, common-variant, and integrated polygenic risk scores for selected disease endpoints.** For each disease, the left panel shows the number of cases as a function of PRS rank for the coding rvPRS, noncoding rvPRS, combined rvPRS, cvPRS, and unified PRS integrating rvPRSs and cvPRSs. The vertical dashed gray line indicates the top 1% threshold of the PRS distribution. The middle panel shows genes and functional annotations enriched among individuals identified by the coding rvPRS and cvPRS. The right panel shows genes and functional annotations enriched among individuals identified by the noncoding rvPRS and cvPRS.

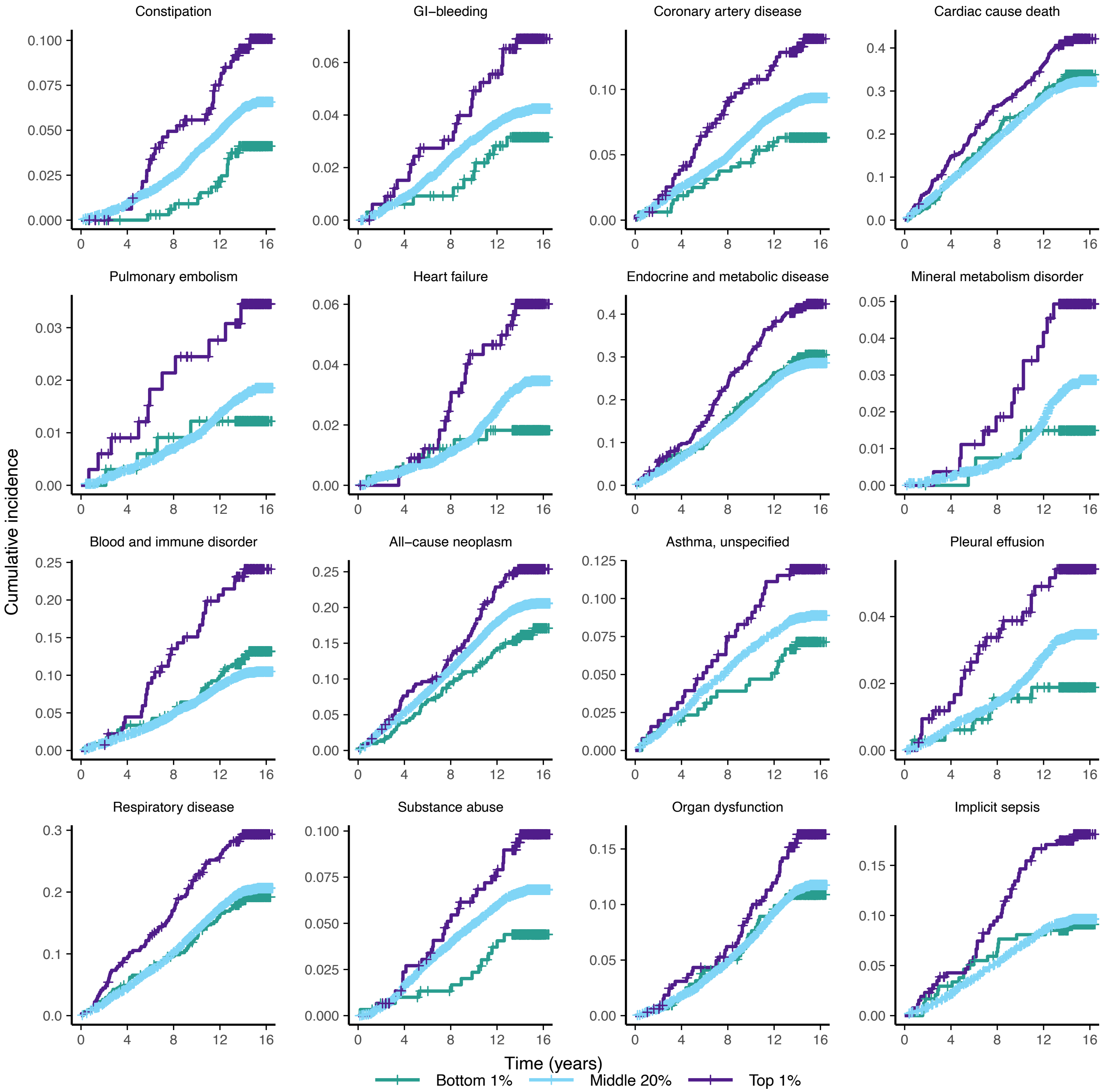


**Supplementary Figure 8: Additional examples of coding rare-variant polygenic risk scores predicting disease onset and stratifying disease trajectories over 15 years of follow-up.** Kaplan-Meier curves show the onset of representative diseases, stratified by the top 1%, middle 20%, and bottom 1% of the coding rvPRS distribution.

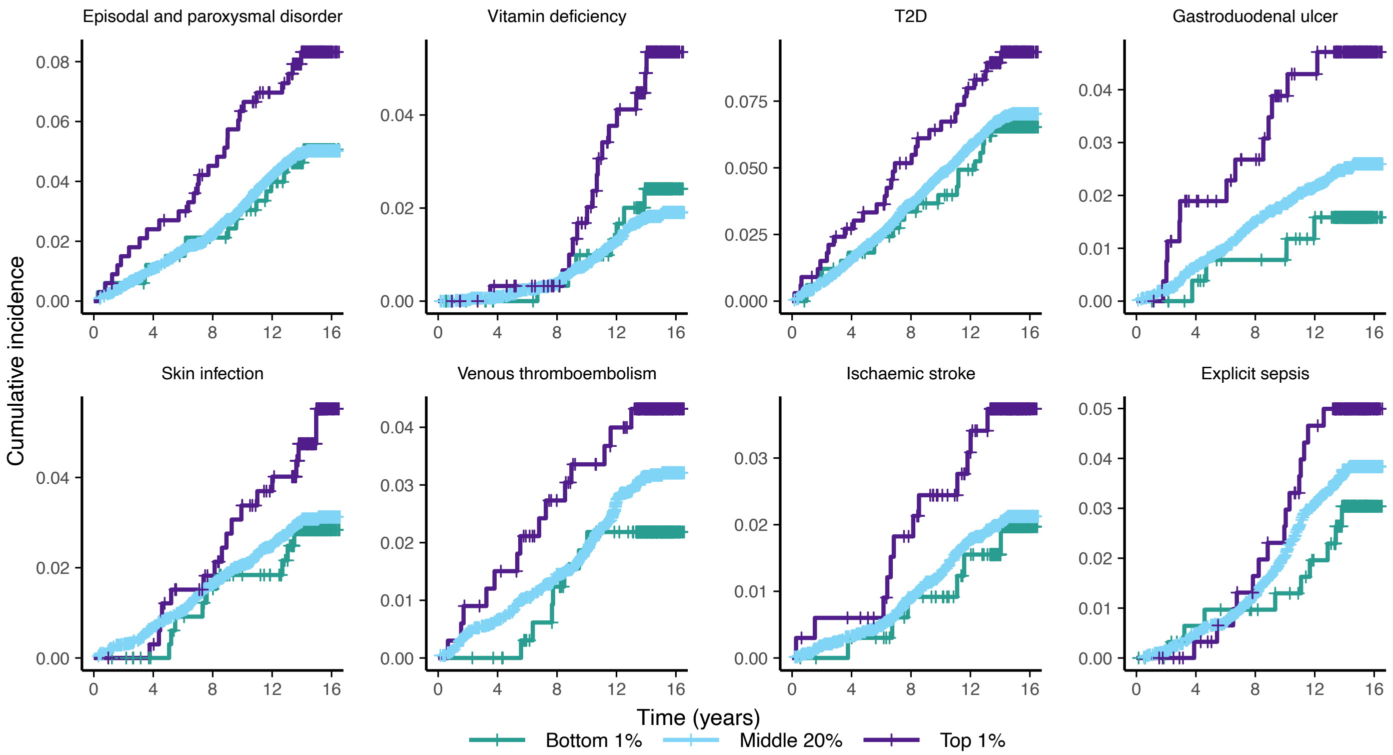


**Supplementary Figure 9: Additional examples of noncoding rare-variant polygenic risk scores predicting disease onset and stratifying disease trajectories over 15 years of follow-up.** Kaplan-Meier curves show the onset of representative diseases, stratified by the top 1%, middle 20%, and bottom 1% of the noncoding rvPRS distribution.

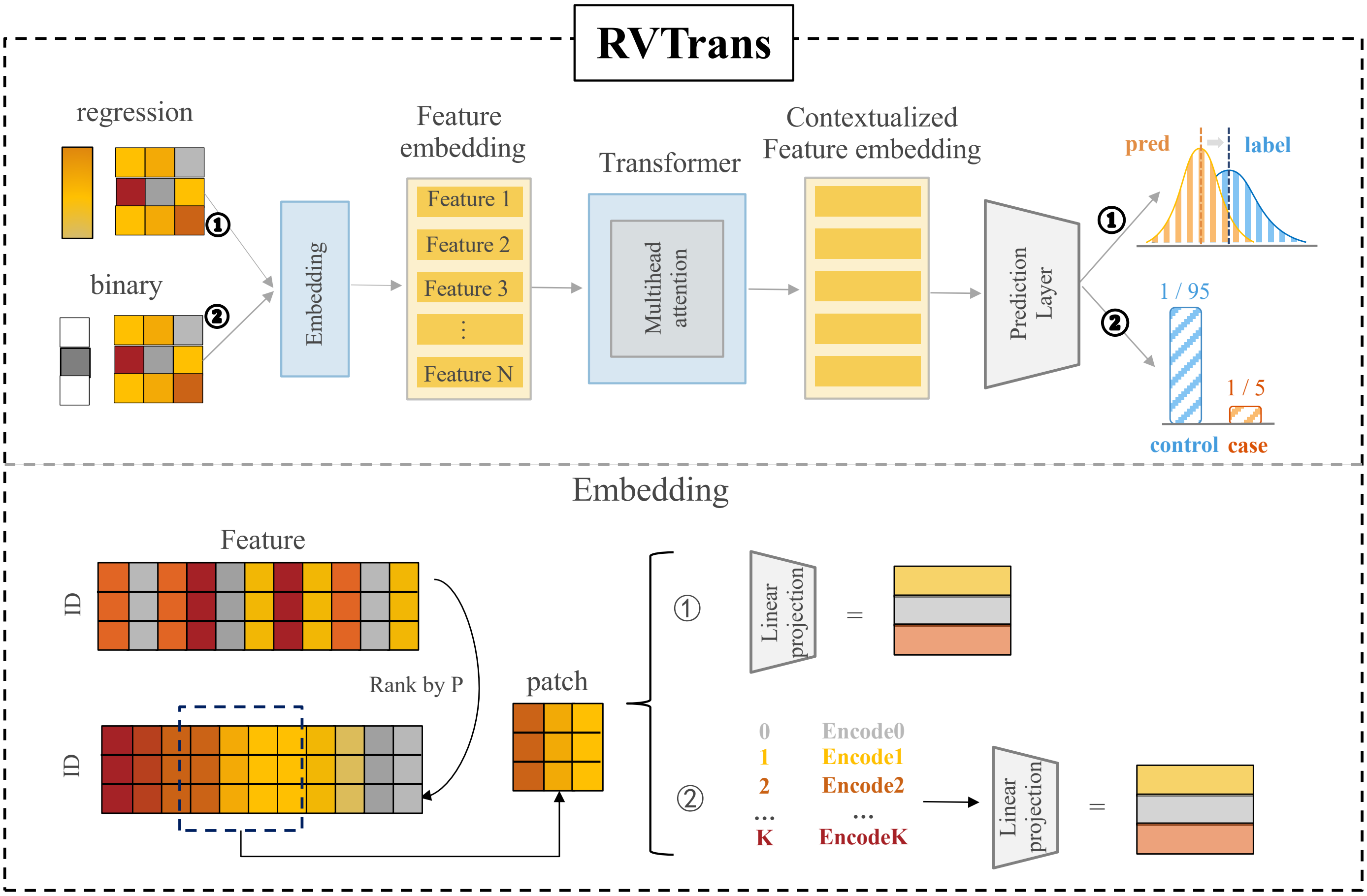


**Supplementary Figure 10: Architecture of RVTrans**. RVTrans uses a feature-ranking mechanism to segment input features into patches, encodes them as embeddings, and applies self-attention mechanisms to generate contextualized representations for prediction. The model uses HL-Gauss loss for quantitative traits and balanced cross-entropy loss for disease endpoints. For quantitative traits, inputs are first mapped through a learnable embedding matrix and then linearly projected into the shared latent space. For disease endpoints, inputs are projected into the same latent space using a one-dimensional convolutional layer that serves as a patch-wise linear projection.
